# COVID-19 Neuropathology at Columbia University Irving Medical Center/New York Presbyterian Hospital

**DOI:** 10.1101/2021.03.16.21253167

**Authors:** Kiran T. Thakur, Emily Happy Miller, Michael D. Glendinning, Osama Al-Dalahmah, Matei A. Banu, Amelia K. Boehme, Alexandra L. Boubour, Samuel L. Bruce, Alexander M. Chong, Jan Claassen, Phyllis L. Faust, Gunnar Hargus, Richard Hickman, Sachin Jambawalikar, Alexander G. Khandji, Carla Y. Kim, Robyn S. Klein, Angela Lignelli-Dipple, Chun-Chieh Lin, Yang Liu, Michael L. Miller, Gul Moonis, Anna S. Nordvig, Jonathan B. Overdevest, Morgan L. Prust, Serge Przedborski, William H. Roth, Allison Soung, Kurenai Tanji, Andrew F. Teich, Dritan Agalliu, Anne-Catrin Uhlemann, James E. Goldman, Peter Canoll

**Affiliations:** Department of Neurology, Vagelos College of Physicians and Surgeons, Columbia University Irving Medical Center, and the New York Presbyterian Hospital, New York, NY, USA; Department of Medicine, Division of Infectious Diseases, Vagelos College of Physicians and Surgeons, Columbia University Irving Medical Center, and the New York Presbyterian Hospital, New York, NY, USA; Department of Pathology and Cell Biology, Division of Neuropathology, Vagelos College of Physicians and Surgeons, Columbia University Irving Medical Center, and the New York Presbyterian Hospital, New York, NY, USA; Department of Neurological Surgery, Vagelos College of Physicians and Surgeons, Columbia University Irving Medical Center, and the New York Presbyterian Hospital, New York, NY, USA; Department of Radiology, Vagelos College of Physicians and Surgeons, Columbia University Irving Medical Center, and the New York Presbyterian Hospital, New York, NY, USA; Department of Otolaryngology, Vagelos College of Physicians and Surgeons, Columbia University Irving Medical Center, and the New York Presbyterian Hospital, New York, NY, USA; Department of Neuroscience Vagelos College of Physicians and Surgeons, Columbia University Irving Medical Center, and the New York Presbyterian Hospital, New York, NY, USA; Departments of Medicine, Pathology & Immunology, Neurosciences, Washington University School of Medicine, St. Louis, MO, USA; Department of Pathology and Laboratory Medicine, Dartmouth-Hitchcock Medical Center, Lebanon, NH

**Keywords:** COVID-19, SARS-CoV-2, neuropathology, microglia activation, microglial nodules

## Abstract

Many patients with SARS-CoV-2 infection develop neurological signs and symptoms, though, to date, little evidence exists that primary infection of the brain is a significant contributing factor. We present the clinical, neuropathological, and molecular findings of 41 consecutive patients with SARS-CoV-2 infections who died and underwent autopsy in our medical center. The mean age was 74 years (38-97 years), 27 patients (66%) were male and 34 (83%) were of Hispanic/Latinx ethnicity. Twenty-four patients (59%) were admitted to the intensive care unit (ICU). Hospital-associated complications were common, including 8 (20%) with deep vein thrombosis/pulmonary embolism (DVT/PE), 7 (17%) patients with acute kidney injury requiring dialysis, and 10 (24%) with positive blood cultures during admission. Eight (20%) patients died within 24 hours of hospital admission, while 11 (27%) died more than 4 weeks after hospital admission. Neuropathological examination of 20-30 areas from each brain revealed hypoxic/ischemic changes in all brains, both global and focal; large and small infarcts, many of which appeared hemorrhagic; and microglial activation with microglial nodules accompanied by neuronophagia, most prominently in the brainstem. We observed sparse T lymphocyte accumulation in either perivascular regions or in the brain parenchyma. Many brains contained atherosclerosis of large arteries and arteriolosclerosis, though none had evidence of vasculitis. Eighteen (44%) contained pathologies of neurodegenerative diseases, not unexpected given the age range of our patients. We examined multiple fresh frozen and fixed tissues from 28 brains for the presence of viral RNA and protein, using quantitative reverse-transcriptase PCR (qRT-PCR), RNAscope, and immunocytochemistry with primers, probes, and antibodies directed against the spike and nucleocapsid regions. qRT-PCR revealed low to very low, but detectable, viral RNA levels in the majority of brains, although they were far lower than those in nasal epithelia. RNAscope and immunocytochemistry failed to detect viral RNA or protein in brains. Our findings indicate that the levels of detectable virus in COVID-19 brains are very low and do not correlate with the histopathological alterations. These findings suggest that microglial activation, microglial nodules and neuronophagia, observed in the majority of brains, do not result from direct viral infection of brain parenchyma, but rather likely from systemic inflammation, perhaps with synergistic contribution from hypoxia/ischemia. Further studies are needed to define whether these pathologies, if present in patients who survive COVID-19, might contribute to chronic neurological problems.

## Introduction

Neurological manifestations of SARS-CoV-2 infection are increasingly recognized^1^ and include seizures^2,3^, cerebrovascular accidents^4,5,6,7,8^, encephalopathy^7^, isolated cases of acute necrotizing hemorrhagic encephalopathy^9^, and Guillain-Barré Syndrome^10^. Many of the neurological conditions seen in the context of SARS-CoV-2 infection are associated with systemic effects, including multiorgan damage, hypercoagulability and a proinflammatory state^11,12,13,14^. Whether SARS-CoV-2 directly infects the brain remains controversial. Related β-coronaviruses such as SARS and MERS show neuroinvasive potential^15, 16,1,17^ and clinicians have reported cases of meningoencephalitis in COVID-19 patients, without neuropathological confirmation^18,19,20,21,3,22,23,16^. Further characterization of the neuropathological and molecular findings in COVID-19 brains is needed to understand the pathophysiological mechanisms underlying the neurological conditions seen in the context of COVID-19.

A few autopsy studies have reported the neuropathological findings of patients with SARS-CoV-2 infections (Supplementary Table 1), including hypoxic changes, vascular lesions, demyelinating pathology resembling acute disseminated encephalomyelitis (ADEM)^24^, reactive astrocytosis and microgliosis^25^ and cerebral hemorrhage or hemorrhagic suffusion^26^. Viral RNA by quantitative reverse transcriptase-PCR (qRT-PCR) has been detected at low levels in some cases ^27,28,29^, but evidence directly linking virus to the COVID-associated neuropathology is controversial. We present the neuropathological findings of 41 consecutive patients with histories of confirmed SARS-CoV-2 infection, investigating the clinical and pathological characteristics.

**Table 1.**
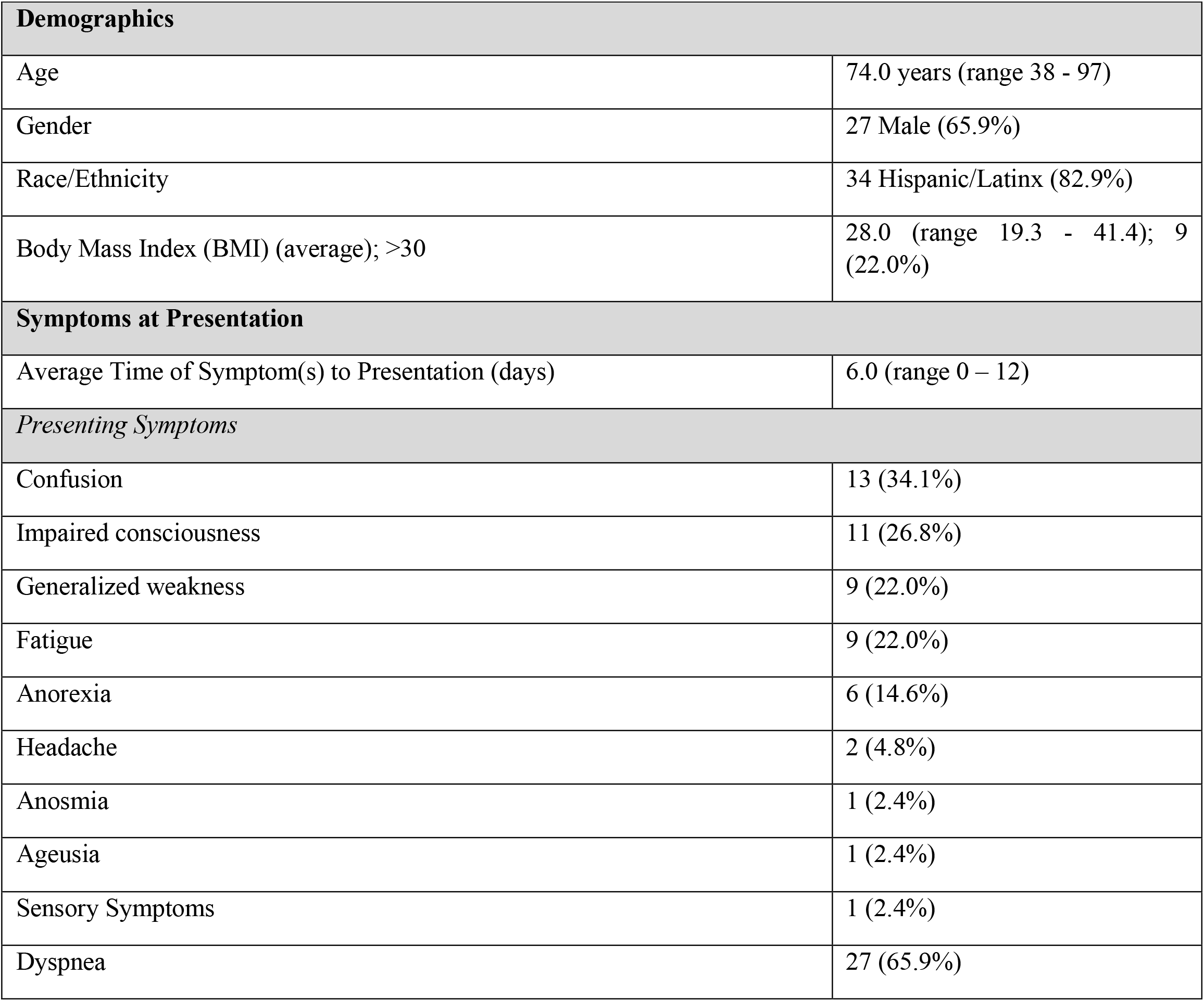
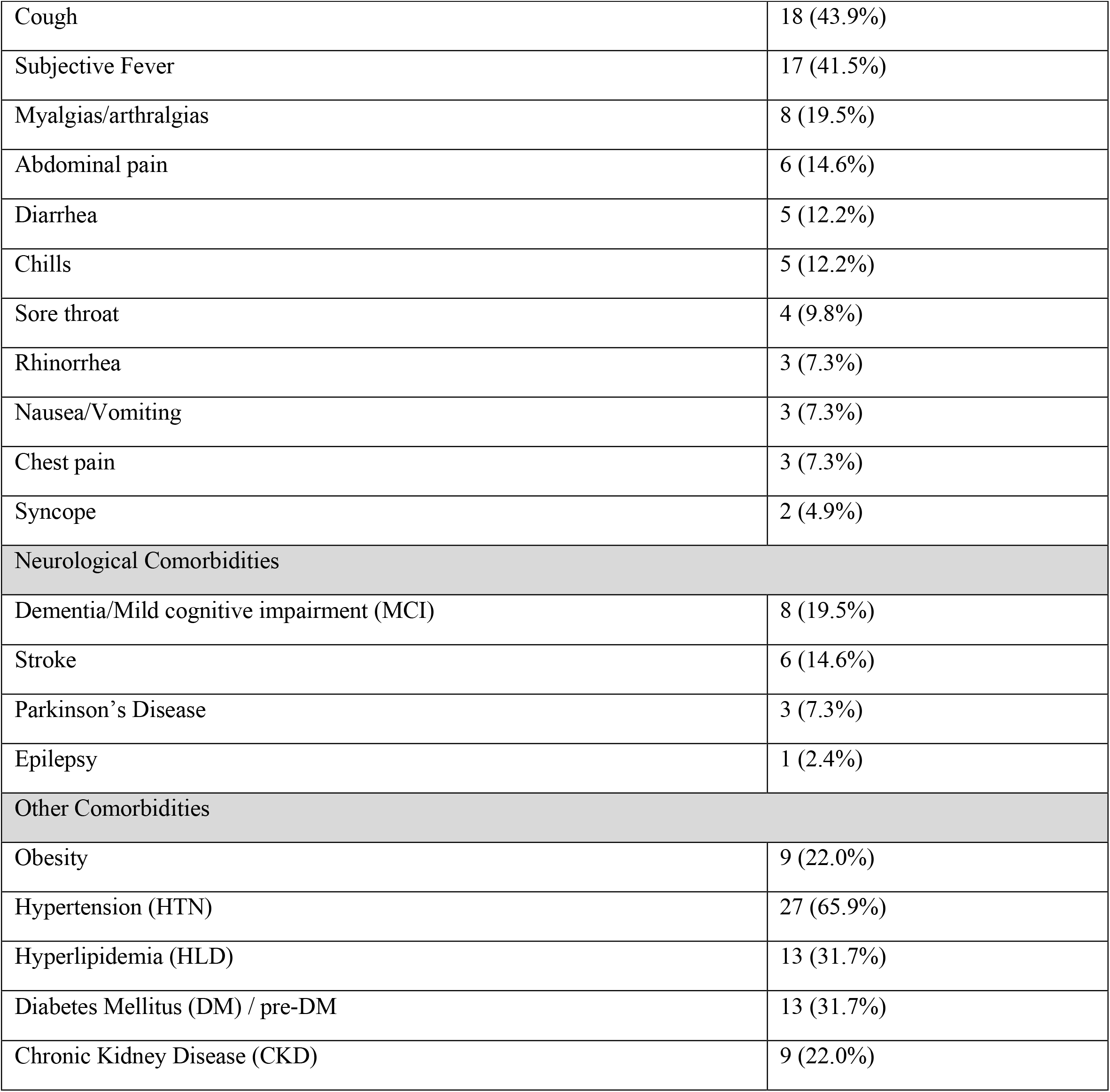
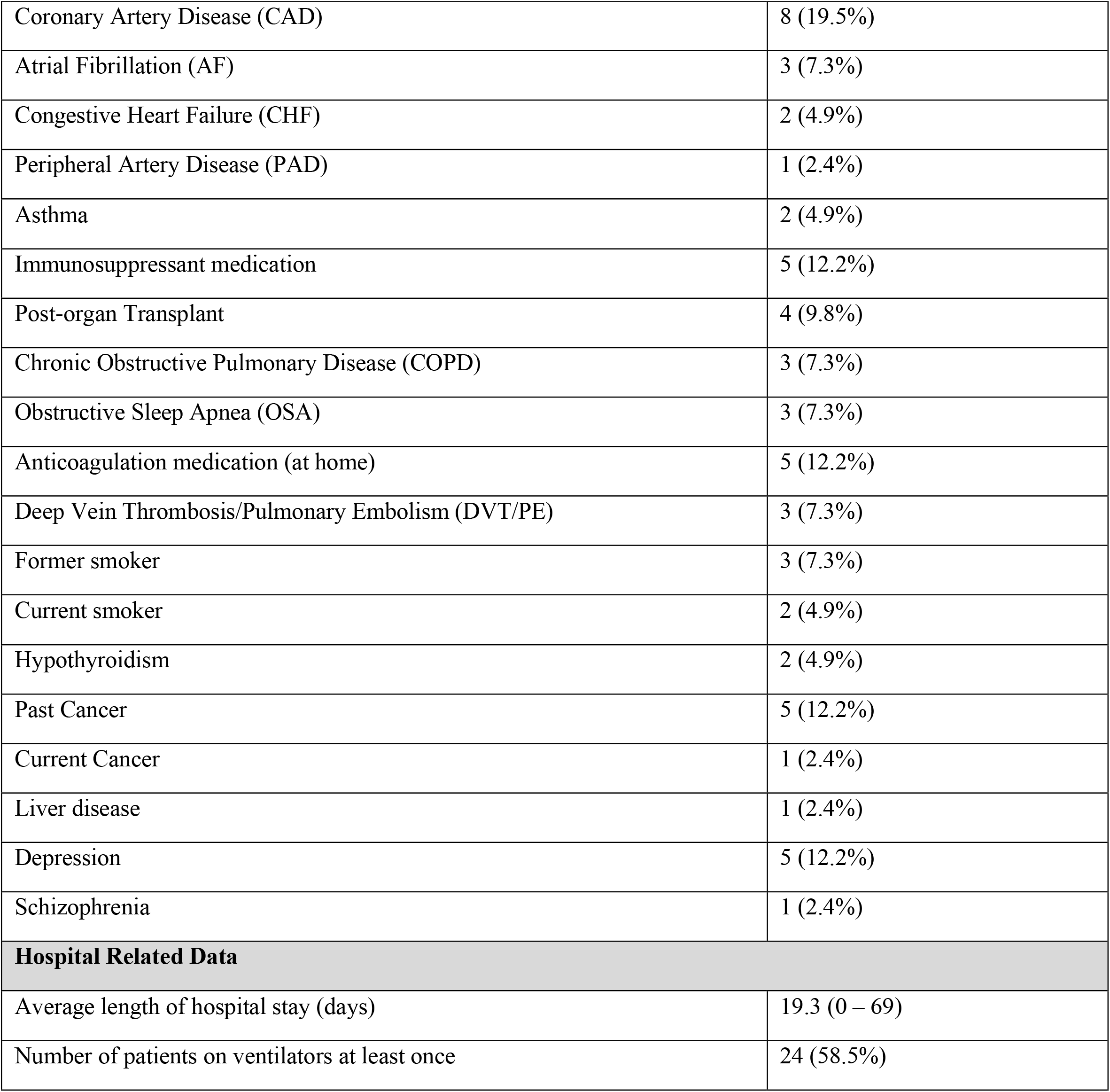
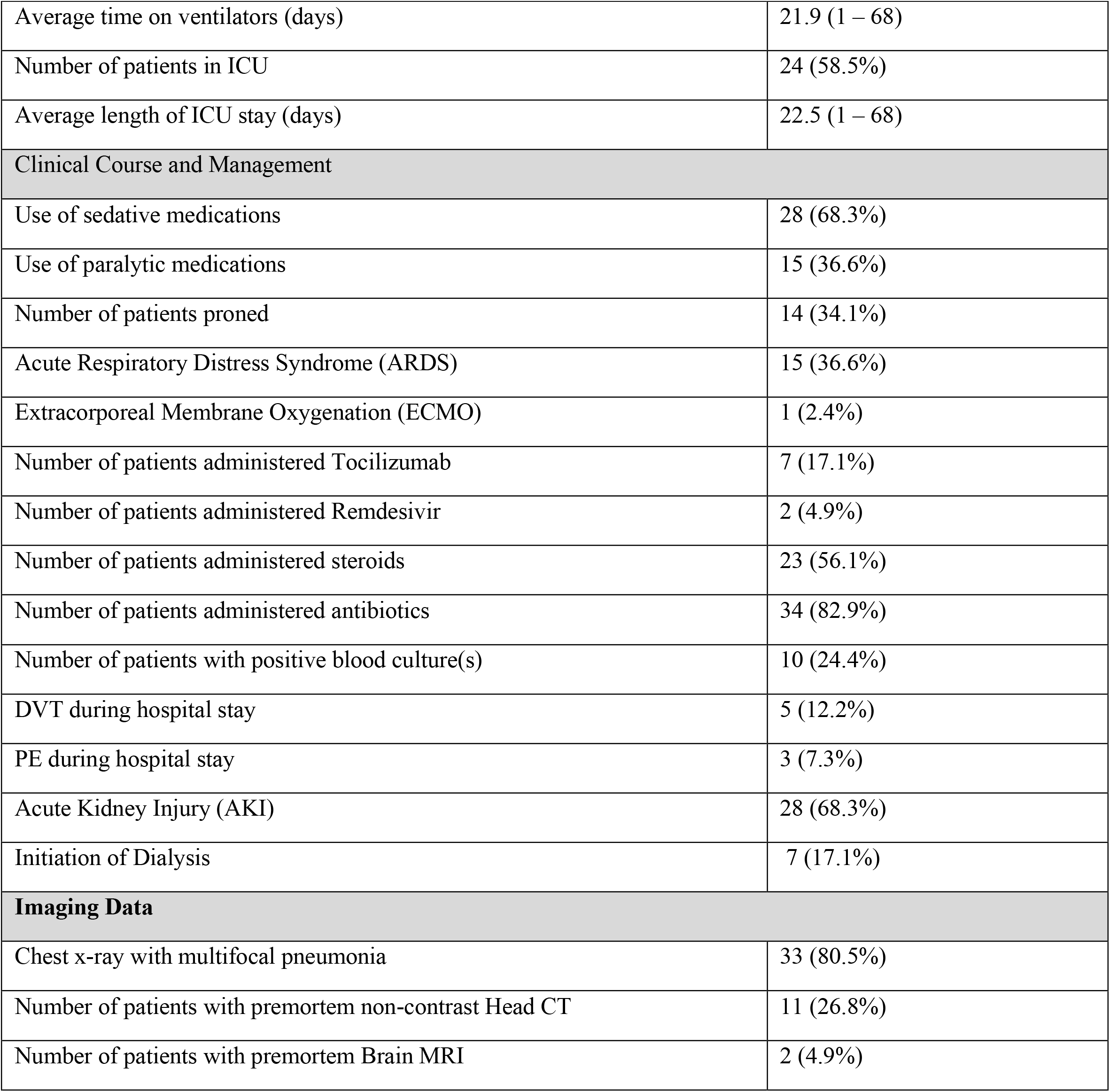
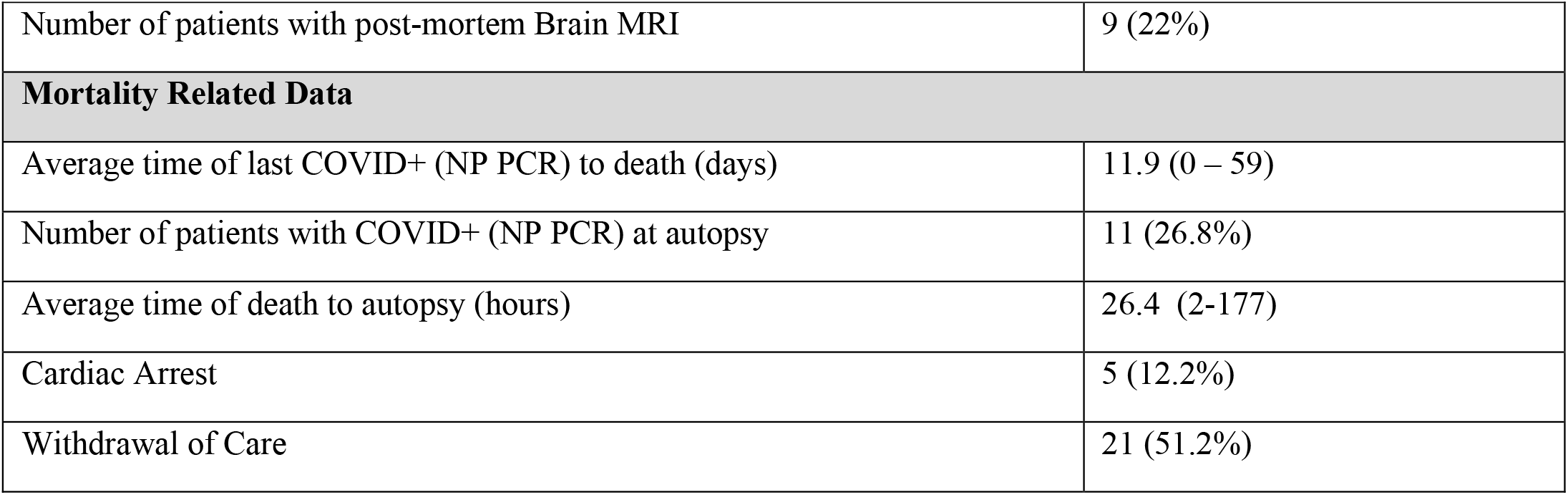
Patient Demographics, Inpatient Characteristics, Complications and Mortality related data

| **Demographics** |  |
| --- | --- |
| Age | 74.0 years (range 38 - 97) |
| Gender | 27 Male (65.9%) |
| Race/Ethnicity | 34 Hispanic/Latinx (82.9%) |
| Body Mass Index (BMI) (average); >30 | 28.0 (range 19.3 - 41.4); 9 (22.0%) |
| **Symptoms at Presentation** |  |
| Average Time of Symptom(s) to Presentation (days) | 6.0 (range 0 – 12) |
| *Presenting Symptoms* |  |
| Confusion | 13 (34.1%) |
| Impaired consciousness | 11 (26.8%) |
| Generalized weakness | 9 (22.0%) |
| Fatigue | 9 (22.0%) |
| Anorexia | 6 (14.6%) |
| Headache | 2 (4.8%) |
| Anosmia | 1 (2.4%) |
| Ageusia | 1 (2.4%) |
| Sensory Symptoms | 1 (2.4%) |
| Dyspnea | 27 (65.9%) |
| Cough | 18 (43.9%) |
| Subjective Fever | 17 (41.5%) |
| Myalgias/arthralgias | 8 (19.5%) |
| Abdominal pain | 6 (14.6%) |
| Diarrhea | 5 (12.2%) |
| Chills | 5 (12.2%) |
| Sore throat | 4 (9.8%) |
| Rhinorrhea | 3 (7.3%) |
| Nausea/Vomiting | 3 (7.3%) |
| Chest pain | 3 (7.3%) |
| Syncope | 2 (4.9%) |
| Neurological Comorbidities |  |
| Dementia/Mild cognitive impairment (MCI) | 8 (19.5%) |
| Stroke | 6 (14.6%) |
| Parkinson's Disease | 3 (7.3%) |
| Epilepsy | 1 (2.4%) |
| Other Comorbidities |  |
| Obesity | 9 (22.0%) |
| Hypertension (HTN) | 27 (65.9%) |
| Hyperlipidemia (HLD) | 13 (31.7%) |
| Diabetes Mellitus (DM) / pre-DM | 13 (31.7%) |
| Chronic Kidney Disease (CKD) | 9 (22.0%) |
| Coronary Artery Disease (CAD) | 8 (19.5%) |
| Atrial Fibrillation (AF) | 3 (7.3%) |
| Congestive Heart Failure (CHF) | 2 (4.9%) |
| Peripheral Artery Disease (PAD) | 1 (2.4%) |
| Asthma | 2 (4.9%) |
| Immunosuppressant medication | 5 (12.2%) |
| Post-organ Transplant | 4 (9.8%) |
| Chronic Obstructive Pulmonary Disease (COPD) | 3 (7.3%) |
| Obstructive Sleep Apnea (OSA) | 3 (7.3%) |
| Anticoagulation medication (at home) | 5 (12.2%) |
| Deep Vein Thrombosis/Pulmonary Embolism (DVT/PE) | 3 (7.3%) |
| Former smoker | 3 (7.3%) |
| Current smoker | 2 (4.9%) |
| Hypothyroidism | 2 (4.9%) |
| Past Cancer | 5 (12.2%) |
| Current Cancer | 1 (2.4%) |
| Liver disease | 1 (2.4%) |
| Depression | 5 (12.2%) |
| Schizophrenia | 1 (2.4%) |
| **Hospital Related Data** |  |
| Average length of hospital stay (days) | 19.3 (0 – 69) |
| Number of patients on ventilators at least once | 24 (58.5%) |
| Average time on ventilators (days) | 21.9 (1 – 68) |
| Number of patients in ICU | 24 (58.5%) |
| Average length of ICU stay (days) | 22.5 (1 – 68) |
| **Clinical Course and Management** |  |
| Use of sedative medications | 28 (68.3%) |
| Use of paralytic medications | 15 (36.6%) |
| Number of patients prone | 14 (34.1%) |
| Acute Respiratory Distress Syndrome (ARDS) | 15 (36.6%) |
| Extracorporeal Membrane Oxygenation (ECMO) | 1 (2.4%) |
| Number of patients administered Tocilizumab | 7 (17.1%) |
| Number of patients administered Remdesivir | 2 (4.9%) |
| Number of patients administered steroids | 23 (56.1%) |
| Number of patients administered antibiotics | 34 (82.9%) |
| Number of patients with positive blood culture(s) | 10 (24.4%) |
| DVT during hospital stay | 5 (12.2%) |
| PE during hospital stay | 3 (7.3%) |
| Acute Kidney Injury (AKI) | 28 (68.3%) |
| Initiation of Dialysis | 7 (17.1%) |
| **Imaging Data** |  |
| Chest x-ray with multifocal pneumonia | 33 (80.5%) |
| Number of patients with premortem non-contrast Head CT | 11 (26.8%) |
| Number of patients with premortem Brain MRI | 2 (4.9%) |
| Number of patients with post-mortem Brain MRI | 9 (22%) |
| **Mortality Related Data** |  |
| Average time of last COVID+ (NP PCR) to death (days) | 11.9 (0 – 59) |
| Number of patients with COVID+ (NP PCR) at autopsy | 11 (26.8%) |
| Average time of death to autopsy (hours) | 26.4 (2-177) |
| Cardiac Arrest | 5 (12.2%) |
| Withdrawal of Care | 21 (51.2%) |

## Methods

This study was approved by the Columbia University Irving Medical Center (CUIMC) Institutional Review Board and is in line with the Declaration of Helsinki. The requirement for written informed consent for chart review was waived as the study design was deemed to be no more than minimal risk. The Institutional Review Board approved this study (AAAS9987) on May 4, 2020. Consent for autopsy was obtained from patient surrogates through standardized consenting procedures via telephone, given that no visitors were allowed in hospital during the study time. We included all consecutive autopsies performed between March through June, 2020, meeting the Center for Disease Control and Prevention case definition for definitive COVID-19 infection (https://www.n.cdc.gov/nndss/conditions/coronavirus-disease-2019-covid-19/case-definition/2020/). Clinical data including demographics, clinical, laboratory, radiographic, and treatment data were obtained retrospectively from electronic medical record (EMR) review. All laboratory tests, neuroradiology assessments, and interventions were performed at the discretion of the treating physicians.

### Autopsy procedures

All autopsies were conducted in a negative pressure room. The brain with the attached uppermost spinal cord was removed after the general autopsy using a bone saw with a vacuum attachment to minimize personnel exposure to bone dust. Sections (up to approximately 3 cm in greatest dimension) from the olfactory bulb/gyrus rectus, superior frontal gyrus, mesial temporal lobe with the anterior hippocampus and amygdala, cerebellum and medulla oblongata were placed in clean plastic bags, sealed, marked, and frozen on dry ice and stored at −80°C in the Columbia University BioBank laboratory dedicated to storing and processing COVID-19 autopsy tissues. After removing tissue for freezing, each brain was fixed in 10% buffered formalin for 10 days. After brain removal, parallel longitudinal cuts were made in the medial anterior cranial fossa to circumferentially excise the cribriform plate of the ethmoid bones along with underlying olfactory and nasal epithelium. A small piece of nasal epithelial tissue was sampled for qRT-PCR, and the remainder divided for freezing and for fixation in 10% formalin.

### Tissue processing

After approximately 10 days in formalin fixation, brains were externally examined, the cerebral hemispheres were sliced in the coronal plane, the brainstem in the transverse plane, and the cerebellum in the sagittal plane. An extensive set of tissue blocks were processed for paraffin embedding in cassettes, and included olfactory bulb/tracts with adjacent gyrus rectus, superior frontal cortex, striatum at the level of the globus pallidus, anterior thalamus, anterior hippocampus, hippocampal formation at the level of the lateral geniculate body, amygdala, striatum at the level of the nucleus accumbens, calcarine cortex, cerebellum with dentate nucleus, corpus callosum (genu), corpus callosum (body with cingulate cortex), corpus callosum (splenium), rostral midbrain, caudal midbrain, rostral pons, caudal pons, four sequential slices of the medulla, cervical spinal cord, pituitary gland, pineal gland, choroid plexus from the lateral ventricle, nasal epithelium, and vessels of the Circle of Willis. Additional blocks were obtained depending on gross pathology, such as infarcts or hemorrhages.

### Tissue staining

Sections of paraffin blocks were cut at 7 μm thickness and stained with hematoxylin and eosin. As routine measures, immunostaining for CD3, CD68, and glial fibrillary acidic protein (GFAP) were performed on sections of olfactory bulb with adjacent gyrus rectus, temporal lobe with hippocampus, pons, medulla, and cerebellum. Immunostaining for SARS-CoV-2 nucleocapsid (N) protein (Sino Biological©; Cat #40143-R001) was performed on sections of nasal epithelium, olfactory bulb, and medulla. Other sections were stained with these antibodies when appropriate. Immunostaining for the majority of cases was conducted in the Department of Pathology Immunohistochemistry Core Laboratory using routine protocols with the Leica™ Bond auto-stainer. All antibodies used are available for clinical diagnostics, including HSV1 (Cell Marque #361A-ASR)

In a subset of cases, additional immunoperoxidase staining was performed on paraffin sections of the pons and choroid plexus from the lateral ventricle in a subset of patients as described^30^. Sequential sections were immunostained for CLAUDIN5 (ThermoScientific 1:200), CD3 (Novus Biological, 1:100), CD31 (Dako, 1:50), CD68 (Novus Biologicals, 1:200), Iba1 (WAKO, 1:250), Collagen IV, (Abcam, 1:300), Laminin (Sigma; 1:60), VCAM-1 (Abcam; 1:250), and ZO-1 (ThermoScientific 1:200). Images were acquired with a Zeiss Axioimager using a color camera and 10X or 20X objective.

### Tissue analysis

Each brain was examined by at least 2 board-certified neuropathologists, and the key pathological findings were reviewed and discussed at a Divisional conference (which includes 8 board-certified neuropathologists). The findings reflect consensus of our group.

### Detection of SARS-CoV-2 by qRT-PCR in brain autopsy tissues

qRT-PCR was performed on 125 brain tissue samples from 25 autopsy cases. Samples were taken from different anatomic locations including nasal epithelium (N=21), olfactory bulb (N=25), superior frontal gyrus (N=7), temporal lobe (N=25), cerebellum (N=23), and medulla oblongata (N=24). The samples were obtained from either fresh or frozen sections and immediately placed in DNA/RNA shield (ZYMO Research; Cat# R1200-25; Irvine, CA). RNA was extracted using RNeasy Mini Kit (Qiagen) and qRT-PCR was performed using Taqman 4X master mix using SARS-CoV-2 primer/probe sets (IDT) against the N region to detect the presence of virus per CDC recommendations (2019 Novel Coronavirus (2019-nCoV) Real-Time Reverse Transcriptase (RT)–PCR Diagnostic Panel). Each assay included a standard curve to determine the viral load (log10 copies/sample).

### Detection of SARS-CoV-2 by RNAscope^®^ in brain autopsy tissues

The RNAscope^®^ was performed on 21 fresh frozen brain samples. These were from the medulla (N=16) and 4 olfactory bulbs and one cerebellum. For positive controls we used 3 lung specimens from patients that had high SARS-CoV-2 viral loads (cycle threshold (Ct) values from 18-19 for the N region). Five ready-to-use probes were acquired commercially: antisense probe for the N gene of SARS-CoV2 (#846081; cross-reacts with SARS and MERS), antisense probe for the S-gene of SARS-CoV-2 (#848561; unique to SARS-CoV2), negative control antisense probe for the LAT gene of the Herpes simplex virus type 1 (#315651), negative control antisense probe for DapB gene of *B. subtilis* strain SMY (#310043) and positive control antisense probe for the human CLAUDIN-5 gene (#517141; a vascular marker) (ACD Biotechne [ACD], Newark, CA, USA). Probes were comprised of 20-40 ZZ oligo probes, except DapB probe, which is comprised of 10 ZZ oligo probes; sequences of probes are proprietary. RNAscope was performed using a 2.5 HD Red Detection Kit according to the manufacturer’s recommendations (ACD). Briefly, fresh frozen sections were fixed in 4% paraformaldehyde for 10-15 minutes, washed 3x with phosphate buffer saline (PBS) for 5 minutes each wash and dehydrated in graded ethanol (50%, 70%, and 100% ethanol) for 5 minutes. Slides were processed immediately or stored in 100% ethanol at −20°C for up to a week. Slides were dried for 5-10 minutes at room temperature, pretreated with 3% hydrogen peroxide for 10 minutes and Protease IV for 15 minutes (ACD), incubated with probe solution for 2 hours at 40°C in the HybEZ™ oven (ACD). ISH signal was amplified following the manufacturer’s recommendation, except that AMP5 incubation was increased to 45 minutes. Slides were developed in WarpRed chromogen (Biocare Medical) for 10-15 minutes at room temperature, counterstained with hematoxylin (Gill II formulation, Ricca Chemical Company, Arlington, TX) and mounted with Permount (Sigma Aldrich, St Louis, MO). Images were acquired with a Zeiss Axioimager using a color camera and 10X or 20X objective. RNA *in situ* hybridization was also performed in the Department of Pathology clinical laboratory as previously described using 2 probes to the SARS-CoV-2 RNA encoding the spike protein^31^.

### Data Availability

All data is available upon reasonable request.

## Results

### Clinical data

The patients’ mean age was 74 years (range 38-97); 27 patients (66%) were male and 24 (83%) were of Hispanic/Latinx ethnicity. The most frequent presenting symptoms included dyspnea (27/41; 66%), cough (18/41; 44%), and confusion (13/41; 34%) (Table 1, Supplementary Table 2). The average time from symptom onset to hospital presentation was 6 days (0-12), and the average length of hospitalization was 19 days (range 0-69 days) (Figure 1). Three (7%) patients presented in acute cardiopulmonary distress with rapid progression to death. Dementia or mild cognitive impairment (MCI) was the most common neurological co-morbidity (8/41; 20%), identified in EMR review. Chest X-rays on admission showed multifocal pneumonia in 33 (81%) of patients. Twenty-four patients (59%) were admitted to the intensive care unit (ICU). Hospital-associated complications were common, including DVT/PE in 8 (20%) patients, acute renal failure requiring dialysis for 7 (17%), and bacteremia in 10 (24%). Seven (17%) patients had a neurology consult during their hospital admission, and one was admitted to the stroke service. Eight (20%) patients died less than 24 hours after hospital admission, 7 (17%) in less than one week, 15 (37%) in 1-4 weeks, and 11 (27%) patients over 4 weeks after hospital admission (Figure1). The average time of death to autopsy was 26 hours (2-177); 31/41 (76%) of the autopsies were performed within 24 hours. The majority of patients for whom laboratory testing was performed showed elevated inflammatory markers. In the 30 patients with c-reactive protein (CRP) measurements; 12 (40%) patients had peak values over 300 mg/L and 18 (60%) had an average of 186.2 (range 12.9-285.2). Peak interleukin 6 levels were markedly elevated in 26 (96%) of the 27 patients tested (Supplementary Table 3).

**Table 2.** Neuropathology Findings

| **Neuropathology** | **N/Total/(%)** |
| --- | --- |
| Hypoxia | 41/41 (100%) focal to global |
| Infarcts * | 18/41 (43.9%) |
| Hemorrhage | 8/41 (19.5%) |
| Lymphocytic Infiltrates | 38/41 (92.6%) mild |
| Microglial Activation, focal or diffuse | 34/41 (80.5%) |
| Microglial nodules/Neuronophagia | 26/41 (63.4%) |
| Acute Thrombosis | 3/41 (7.3%) |
| Athero- Arteriole-sclerosis | 36/41 (87.8%) mild to severe |
| AD,CAA,PD,PART pathology | 18/41 (43.9%) |
| Herpes Encephalitis | 1/41 (2.4%) |

AD – Alzheimer’s Disease, CAA – Cerebral Amyloid Angiopathy, PD – Parkinson’s Disease, PART – Primary Age-Related Tauopathy
\* Infarcts – 6 brains with chronic infarcts, 12 with acute or subacute infarcts; 7 with microscopic acute or subacute infarcts (not seen grossly); 9 with multiple infarcts, one of them with chronic infarcts; locations: isocortex (9), corpus callosum (5), pons (5), midbrain (1), thalamus (2), caudate (1), putamen (1), hippocampus (2 – both chronic), pituitary (2).

**Figure 1.**
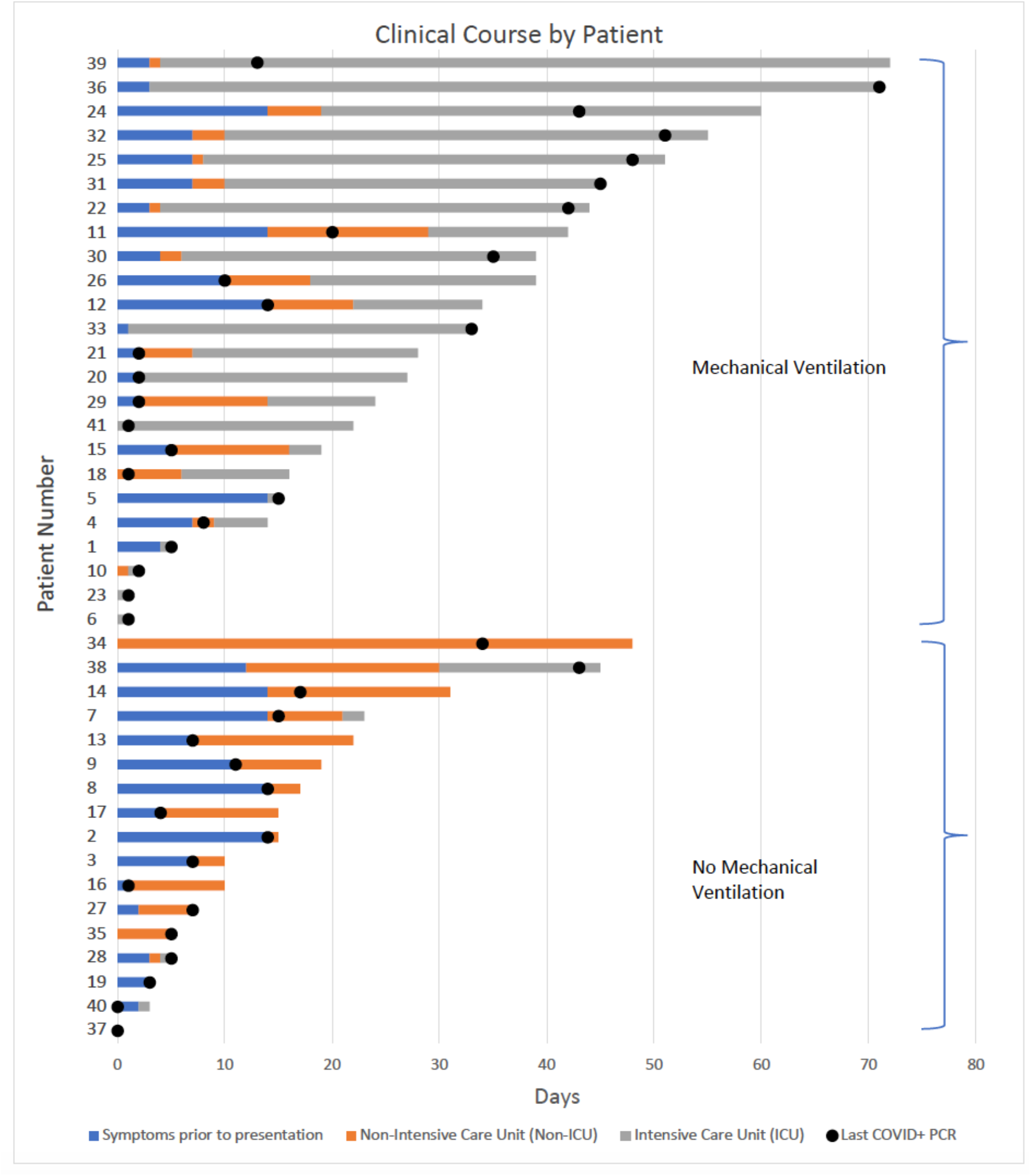
Clinical course of COVID-19 patients by days. Bar graph showing the length of clinical courses of our patients, including symptoms prior to presentation (blue) and days in the hospital before death, either in the ICU (ventilatory support, orange) or not in the ICU (no ventilatory support, gray). Dots represent the last positive qRT-PCR result prior to death. Patients numbered from 1-41 and displayed from shortest to longest clinical course.

### Antemortem and postmortem neuroimaging

Non-contrast head CTs were performed on 11 (27%) patients during their hospital stays and 2 (5%) also underwent a brain MRI (Table 1). Parenchymal hemorrhages/hemorrhagic infarcts were identified in 3 patients and multiple cortical and deep gray nuclei early subacute infarcts in 1 patient. Five patients had diffuse cerebral edema and hypoxemic injury, 3 of whom exhibited concurrent or prior hemorrhages and 1 of these also showed bilateral basal ganglionic petechial hemorrhages. Nine brains (22%) were imaged postmortem: 1 brain with multiple cortical hemorrhagic infarcts and a right basal ganglionic infarct, 1 with mild cortical hemorrhage, 1 with minimal bilateral basal ganglionic hemorrhages, 1 with a right occipital parenchymal hemorrhage, 2 with intraventricular hemorrhages. An example of premortem, postmortem, and corresponding neuropathology of acute hemorrhagic infarcts is shown in Figure 2.

**Figure 2.**
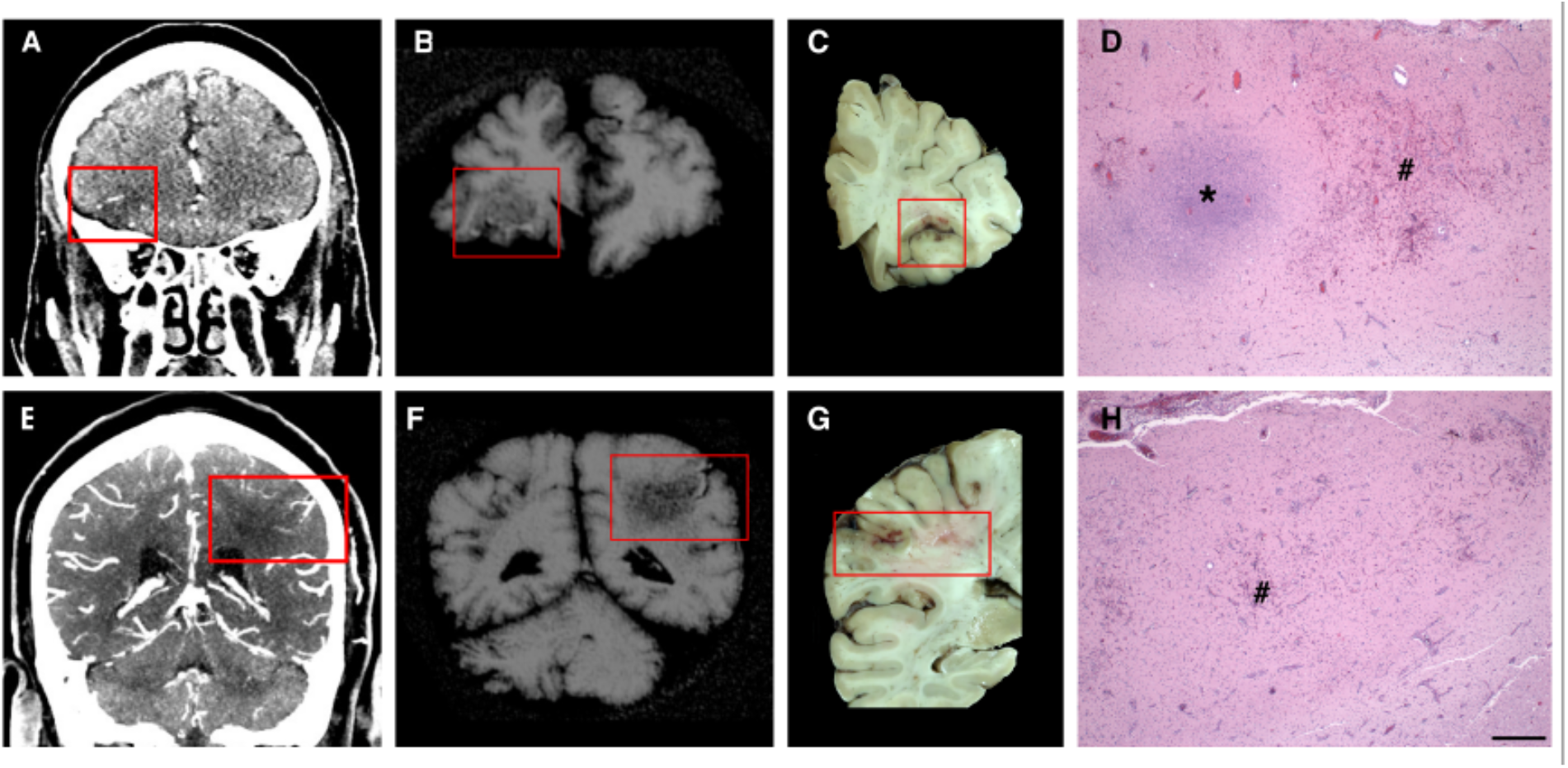
Acute, focal hemorrhagic infarcts in COVID-19 patients. Acute, focal, hemorrhagic infarcts of A-D) right inferior frontal and E-H) left lateral parietal lobes. A,E) premortem CAT scans; B,C) postmortem MRI; C,D) coronal brain slices, and D,H) microscopic images. Boxes represent the corresponding areas on the scans and brain slices. D,H) areas fresh hemorrhage (#) and acute necrosis (*) shown with H&E stain. Scale bar: D, H) 500μm.

### Neuropathological findings

#### Hypoxic/ischemic injury was the most common pathology

All brains contained hypoxic damage, varying from acute to subacute. Acute changes included neuronal shrinkage and eosinophilia with or without neuronal loss, reactive astrocytosis, highlighted with GFAP immunostaining, and subacute hypoxic changes manifested as subacute infarcts with variable macrophage infiltrates, reactive astrocytosis, and neovascularization. These findings were widespread in most brains, but in a few patients (9/41; 22%) were more focal, predominantly involving the isocortex, hippocampus, cerebellum, and/or brainstem (Table 2).

#### Vascular pathology was also common

Eighteen (44%) brains contained infarcts, acute, subacute, or chronic in isocortex, striatum, thalamus, hippocampus, corpus callosum, and brainstem, with 10 (24%) containing one or multiple small infarcts (Table 2). None of the larger infarcts appeared to represent watershed infarcts. Eight (19%) brains contained hemorrhages involving isocortex, white matter, cerebellum, brainstem, and/or the subarachnoid space. These ranged from multifocal, perivenular hemorrhages, to large hemorrhages, the largest in the cerebellum. The majority of hemorrhages appeared to represent ischemic infarcts that became hemorrhagic, evidenced by small hemorrhages adjacent to infarcted tissue (Figure 2A-H).

The majority of brains (36/41; 88%) showed atherosclerotic changes in the Circle of Willis and arteriolosclerosis in smaller intraparenchymal arteries and arterioles. We did not detect vasculitis, defined as fibrinoid necrosis of vessel walls or the destruction of vessels walls with intramural inflammatory cells. Immunostaining for the vascular basement membrane proteins such as Collagen IV and Laminin or the tight junction protein Zona Occludens-1 (ZO1) showed intact capillary and venular walls (Supplementary Figure 1 A-F’). One patient who had disseminated Herpes simplex 1 (HSV-1) infection with CNS involvement (see below), showed a clear loss of vascular basement membrane and ZO-1 immunostaining (Supplementary Figure 1G-I’) and an increase in vascular cell adhesion molecule-1 (VCAM-1; data not shown).

#### Microglial activation was present in the majority of COVID-19 brains

We defined microglial activation by enlargement of cell soma and thickening of processes detected by either Iba1 or CD68 immunostaining. Diffuse microglial activation was present in the majority of the brains (34/41; 81%), variably involving many brain areas (Table 2). Microglia also appeared in clusters (microglial nodules), in over half of the brains (26/41; 63%). Small clusters of CD3^+^/CD8^+^ T cells were associated with prominent microglial nodules in a few cases (Figure 3C,D; Supplementary Figure 2A,B,E,F). Neurons were present in some of these microglial clusters (Figure 3A,B,E,F,G,H,I; Supplementary Figure 2A,B,E,F), representing neuronophagia. The microglial nodules were most prevalent in the brainstem, where they appeared particularly common in the inferior olivary nucleus and the tegmental nuclei of the medulla and pons, including the locus ceruleus, hypoglossal nucleus, dorsal vagal motor nucleus, solitary nucleus, and midline raphe (Figure 3A-G; Supplementary Figure 2A-F). They were also present in the cerebellar deep nuclei (Figure 3H,I) and white matter, although not in the cerebellar cortex, except one patient who had concomitant HSV-1 encephalitis and diffuse microglial activation (Supplementary Figure 2I,J; see below). The microglial nodules were less frequent in the hippocampus (8/41; 20%), where they preferentially localized to the pyramidal cell layer (Figure 3J,K), and in the isocortex (2/41; 5%), and olfactory bulb (2/41; 5%).

**Figure 3.**
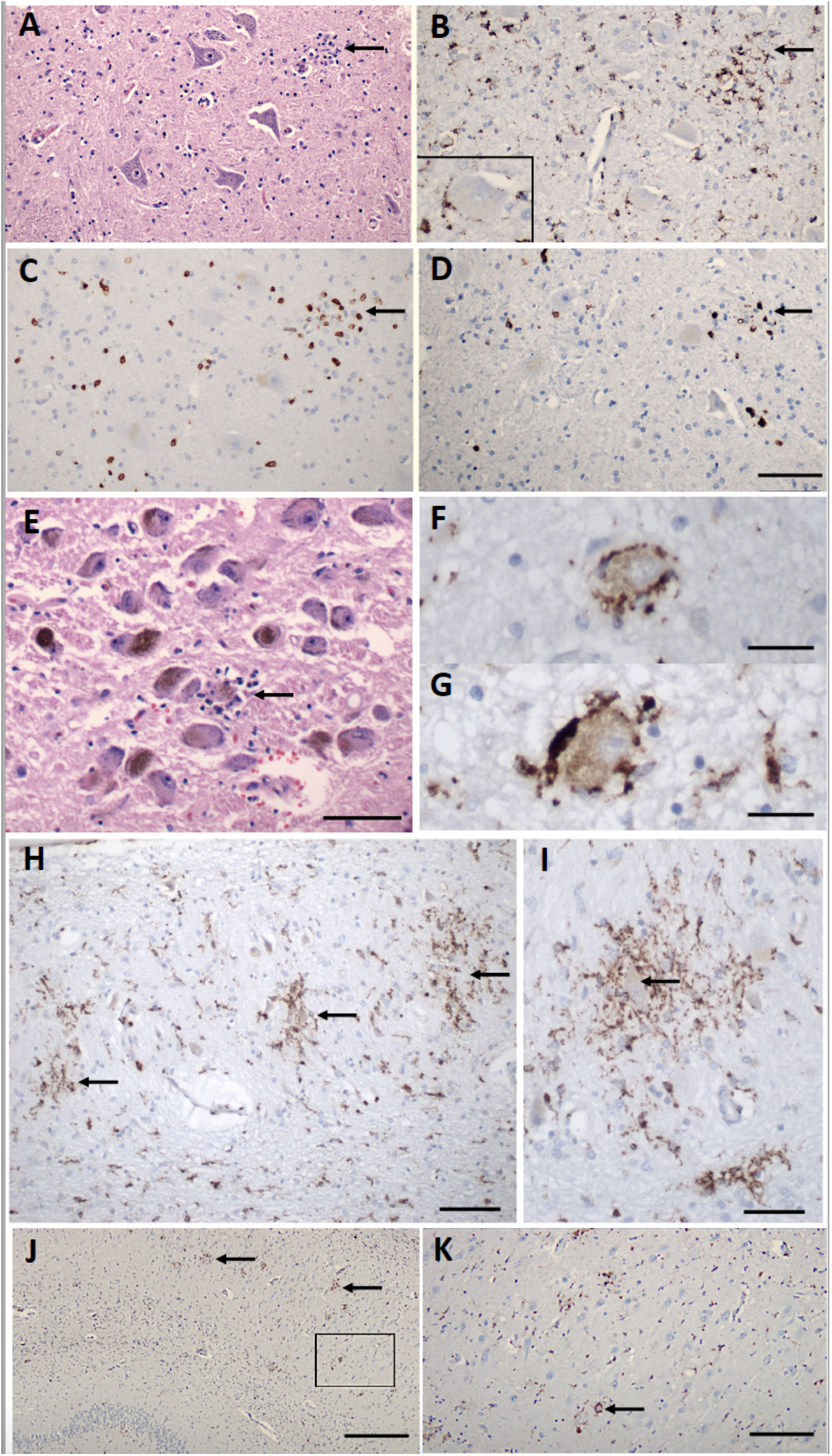
Inflammatory pathology in COVID-19 brains. A) Section of the hypoglossal nucleus shows several motor neurons and a microglial module (arrow). B) An adjacent section stained for CD68, showing clustered microglia in the nodule. Inset – Microglia in close apposition to a hypoglossal neuron (CD68). C) An adjacent section stained for CD3, showing scattered T cells in the tissue and associated with the microglial nodule. D) An adjacent section stained for CD8 showing that many of the T cells are CD8+. E) The locus ceruleus contains a microglial nodule with a degenerating neuron in the center, identified by its residual neuromelanin (arrow). F, G) Neurons of the dorsal motor nucleus of the vagus surrounded by CD68+ microglia. H, I) Microglial nodules in the dentate nucleus (arrows in H), neuron in the middle of a nodule (arrow in I), CD68. J, K Microglial nodules in the pyramidal cell layer of the hippocampus (two marked with arrows), K higher magnification of the box in J (arrow marks neuron surrounded by microglia), CD68. Scale bars: A-D 200μm, E 10μm, F, G 50μm, H, 100μm, I 50μm, J 1mm, K, 250μm.

#### Perivascular lymphocytic inflammation and infiltration into the brain parenchyma was sparse

In 38 (93%) of brains we found scant lymphocytic infiltration, predominantly around blood vessels, with very few CD3^+^ T lymphocytes penetrating into the brain parenchyma and meninges (Figure 3C,D, 4A,A’,C,C’). Immunostaining for CD20 revealed no B lymphocyte infiltration (data not shown). One brain showed very large perivascular and intraparenchymal T cell and macrophage infiltrates concomitant with an HSV-1 infection (Figure 4E, E’; Supplementary Figure 2I,J), providing a stark contrast to the modest level of lymphocytic infiltration seen in COVID-19 brains. We also observed few lymphocytes in the choroid plexi from lateral ventricles of 16 brains, except one with HSV-1 encephalitis that contained prominent T cells and activated microglia (Figure 4B,D,F; Supplementary Figure 2K,L, 5D-F). Consistent with sparse immune cell infiltrations in the CNS of COVID-19 cases, we also found that expression of tight junction proteins in the choroid plexus epithelial cells was intact with the exception of the HSV-1 encephalitis case, where epithelial tight junction staining was completely lost (Supplementary Figure 3).

**Figure 4.**
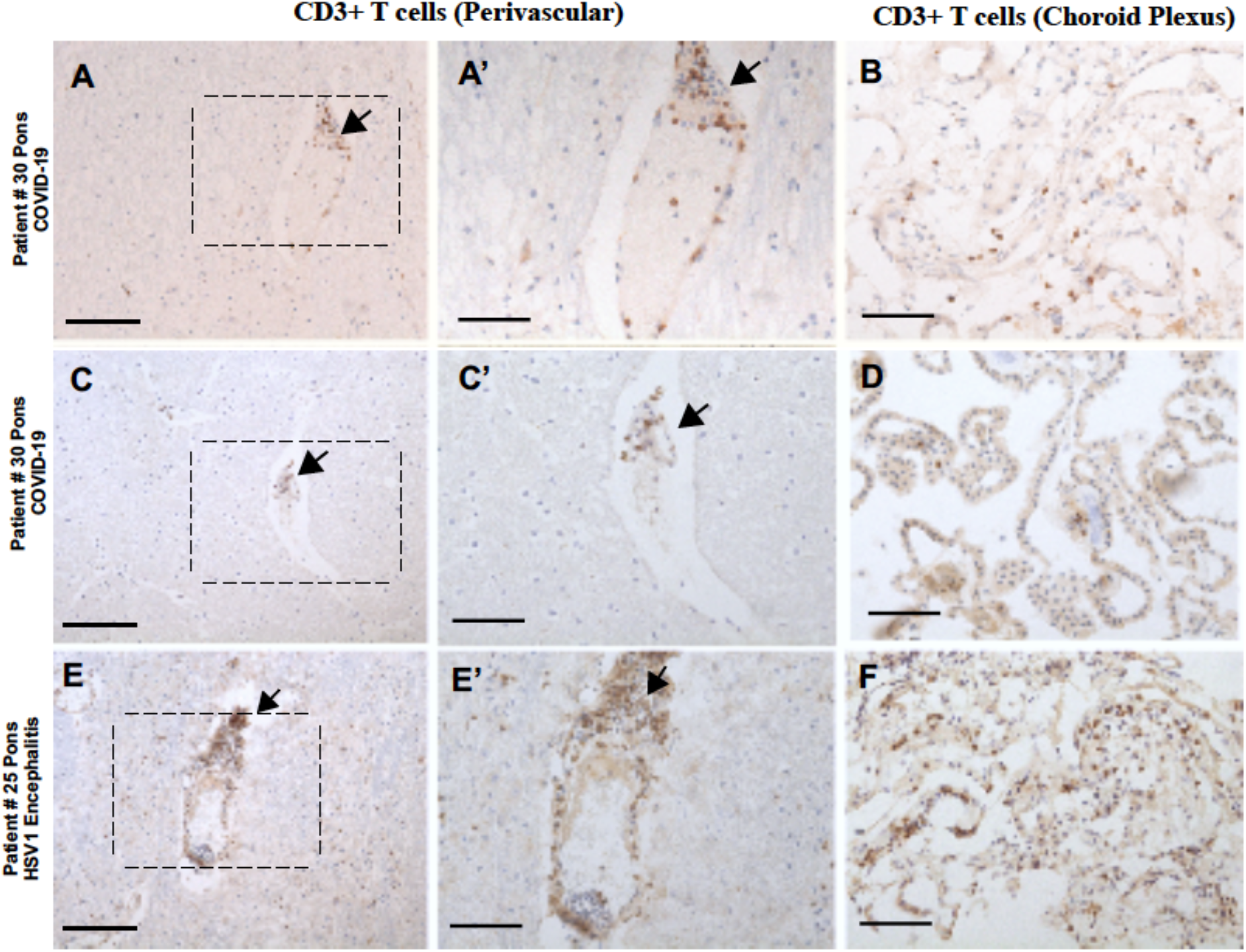
Immunocytochemical staining for CD3+ T cells in COVID-19 brains. A, C) Sparse perivascular CD3+ T cells in the pons of COVID-19 patients #27 and 30. B, D) Sparse CD3+ T cells in the choroid plexus from the lateral ventricle of patients #27 and 30. E,F) Larger CD3+ T cell infiltrates around vessels (E) and in the choroid plexus (F) of the brainstem of patient #25 with HSV-1 encephalitis; A’, B’, E’) are magnified images of boxed areas; arrows represent the same point in the corresponding images. All sections are counterstained with hematoxylin. Scale bars A-F, 100μm, A’, C’, E’ 200μm

#### Many patients showed the pathology of neurodegenerative diseases

Given the ages of our patients and the antemortem histories of dementia/mild cognitive impairment (MCI) and Parkinson’s disease in some patients (Table 2), it was not unexpected that 19 brains contained neurofibrillary tangles, with and without amyloid plaques (17/41, 41.4%), and Lewy bodies of Parkinson’s disease (3/41, 7%). One of these 18 patients had both Alzheimer’s and Parkinson’s pathology. Of note, only 8 had an antemortem diagnosis of dementia/MCI and 3 a history of Parkinson’s disease.

#### Demyelination was not evident

Because of a report of ADEM-like pathology in a COVID-19 patient^24^, we examined all brains for demyelination, but did not find evidence of it. While we cannot rule out the possibility that some of the areas of brains that were not sampled contained foci of demyelination, we note that brains were extensively sampled in 20 - 30 regions for histopathology.

#### Multifocal necrotizing leukoencephalopathy (MNL) in one patient

We found lesions consistent with MNL in the pons in one patient, showing small foci of necrosis, myelin loss, edema, and axonal swellings (Supplementary Figure 4). This woman (age 70-75 years) with hypertension and hypothyroidism, was admitted after one-week of fever, cough and chills in hypoxemic respiratory failure. She developed acute kidney injury requiring renal replacement therapy, severe hypoxemia requiring paralysis and pronation, and multiple other infections (*E. coli* urinary tract infection, methicillin-susceptible *Staphylococcus aureus* pneumonia and recurrent candidiasis in respiratory and urine cultures). The patient’s status acutely worsened with septic shock in the context of *Pseudomonas aeruginosa* ventilator-associated pneumonia and bacteremia.

#### Pathology in the olfactory bulbs was mild

Because of widespread speculation that SARS-CoV-2 may enter the brain via the olfactory route^32^, we examined olfactory bulbs in all patients. We found variable, albeit generally sparse numbers of T cells in the parenchyma, mild to moderate patchy microglial activation and very few microglial nodules, indicating no major histopathology (data not shown).

#### One brain contained HSV-1 encephalitis

One of our patients also had a disseminated HSV-1 infection with CNS involvement, diagnosed only postmortem (patient #22). This woman (age 70-75 years) with hypertension, living independently, developed acute respiratory distress and vasodilatory shock in the context of COVID-19. She developed acute renal failure, *Staphylococcus epidermidis* bacteremia, *Pseudomonas aeruginosa* ventilator-associated pneumonia, and cytomegalovirus viremia during her prolonged hospitalization. Because of her refractory hypoxemia after 44 days of hospitalization, she was palliatively extubated and died within minutes. We found disseminated HSV-1 infection with CNS involvement, proven by immunocytochemistry (Supplementary Figure 5C). In contrast to COVID-19 neuropathology, the brain had prominent lymphocytic infiltrates in the parenchyma, meninges, and choroid plexus, vasculitis, severe microglial activation and microglial nodules (Figure 4E,F; Supplementary Figures 1G-I’, 2I-J, 5). In addition, there was a reduction or loss of Collagen IV and Laminin in the basement membrane of blood vessels loss of ZO1 expression in endothelial cells in proximity to massive T cell infiltrates (Supplementary Figure 1G-I’) and loss of CLAUDIN expression in the choroid plexus epithelial barrier (Supplementary Figure 3D, D’).

### Molecular Findings

#### qRT-PCR detected very low levels of viral RNA in some brains

To determine whether SARS-CoV-2 was present in the brain we conducted qRT-PCR for the N gene in 4 different brain areas in the first 25 patients of this series. We also examined the nasal epithelium for most of these patients as an internal control. Nearly all available nasal epithelium (NE) samples (19/21; 91%) were highly positive for SARS-CoV-2 by qRT-PCR. The median viral copy/sample for NE was 43,840 (Supplementary Table 4). For CNS samples, the proportion of positive samples as well as the number of viral copies was significantly lower (Figure 5, Supplementary Table 4). In 7/25 (28%) of patients, all CNS sites were negative for SARS-CoV-2, whereas 18/25 (72%) of patients had at least one very low but positive CNS site and 9/25 (36%) of patients had multiple very low but positive CNS sites (Figure 5). The cerebellum was most commonly positive (10/23; 44%), followed by samples from the olfactory bulb (10/25; 40%), the temporal lobe (9/25; 36%), and the medulla (8/24; 33). We did find any correlation between detection of viral RNA by qRT-PCR and histopathologic findings discussed above.

**Figure 5.**
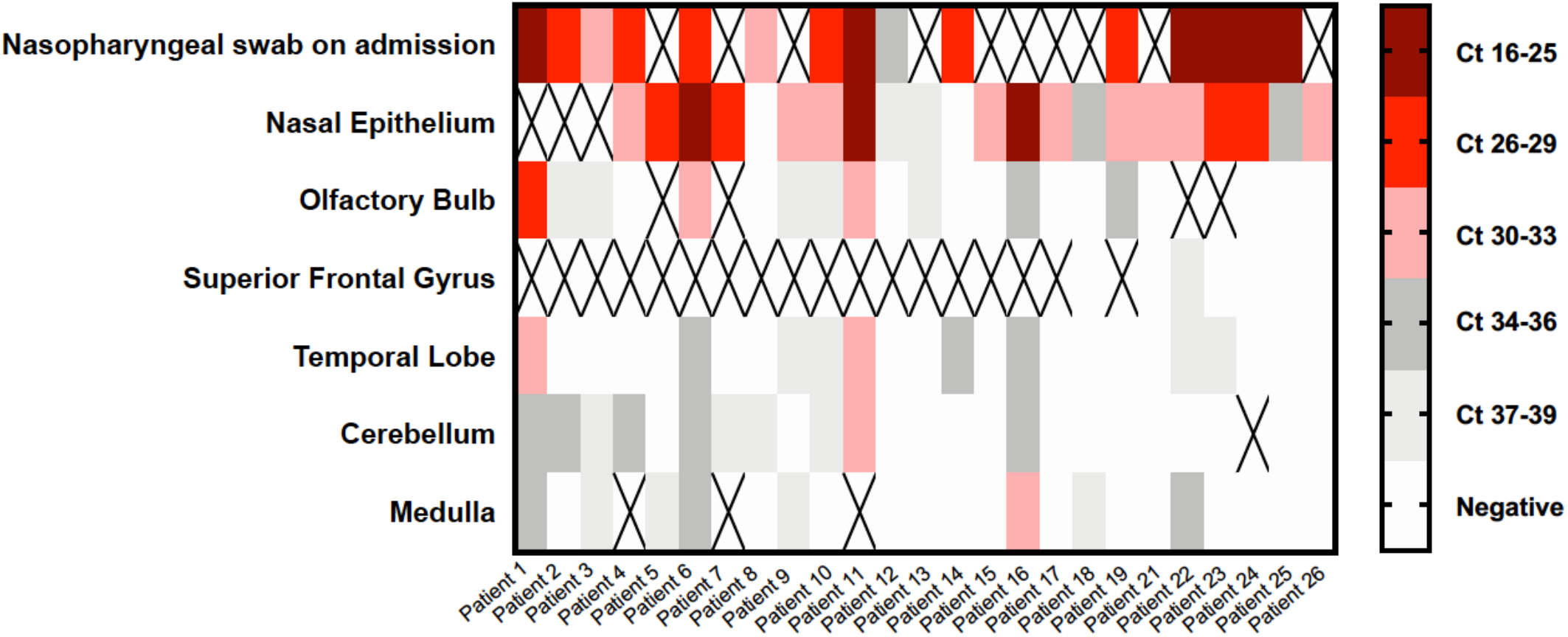
SARS-CoV-2 qRT-PCR results from the nasal epithelia and brains of COVID-19 patients. Heat map of cycle threshold (Ct) values of brain autopsy samples qRT-PCR for detection of SARS-CoV-2. Nasopharyngeal swab at the time of admission and nasal epithelium are included as samples outside of the CNS. Ct values are presented in quintiles based on distribution in samples tested. “X” denotes sample not included for that patient.

#### SARS-CoV-2 mRNA was not detected in brain tissue by RNAscope

The presence of low levels of viral RNA in at least one brain region in 18/25 (72%) of COVID-19 patients raised the question whether the virus is present in the brain or is associated with the vasculature or blood components including infiltrating macrophages. We performed RNAscope on fresh frozen COVID-19 brain sections for 16 cases, including sections from the medulla (N=16), olfactory bulb (N=3) and cerebellum (N=1) (Supplementary Table 5). The brains were selected based on qRT-PCR data to include areas that were either high, low positive, or negative for the viral RNA by RT-PCR. Lung sections from 3 COVID-19 positive cases in a different series that had high CT values (18-19 for the N gene) by RT-PCR were selected as positive controls for the RNA scope. In contrast to the RT-PCR data, we could not detect viral RNA on fresh frozen brain sections using either an antisense probe for the S or N region, or a combination of both probes in cases that were either positive or low positive by RT-PCR (Figure 6A,B,D,E,F,G). However, we could detect abundant RNA for SARS-CoV-2 S or N region in the COVID-19 lung sections (Figure 6C,F,I). As a positive control on brain sections, we detected CLAUDIN-5 mRNA (an endothelial cell marker) in medullas (Figure 6J,J’). Notably, in one case (# 18), we detected viral RNA for the S region in perivascular cells in the adventitia of a large blood vessel outside the medulla, suggesting sporadic infection of blood vessel cells (Figure 6K,K’). Overall, our findings suggest that if SARS-CoV-2 is present in brain tissue, either its levels are very low and below the limits of detection by RNAscope or the virus had already been cleared in some brains.

**Figure 6.**
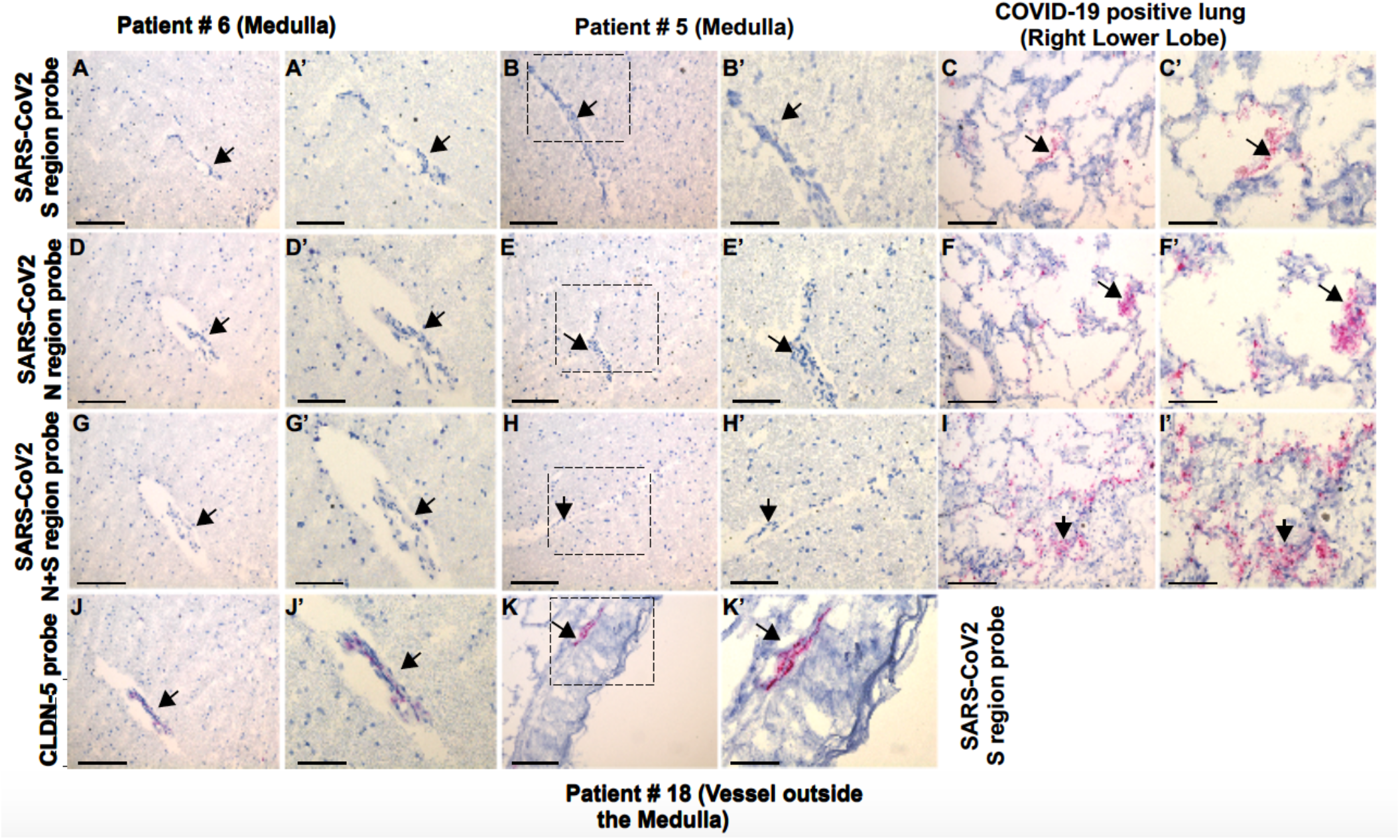
RNAscope results from the brains and lungs of COVID-19 patients. RNAscope with A, B, C) SARS-CoV-2 S region probe on A) medulla patient #6, B) medulla patient #7, C) lung, positive control; D, E, F) SARS-CoV-2 N region probe on D) medulla patient #6, E) medulla patient #7, F) lung, positive control; G, H, I) SARS-CoV-2 N + S region probes on G) medulla patient #6, H) medulla patient #7, I) lung, positive control; J) CLDN-5 probe on medulla patient #6 showing positive signal in endothelial cells; K) SARS-CoV-2 S region probe gives a positive signal in the adventitia of a meningeal vessel outside of medulla patient #18. All sections counterstained with hematoxylin. A’ through K’ are magnified images of boxed areas; arrows represent the same point in the corresponding images. Scale bars: A-K 100μm, A’-K’ 200μm.

#### Immunohistochemistry did not detect viral proteins in COVID-19 brains

We performed immunocytochemistry for SARS-CoV-2 N protein on sections of olfactory bulb and medulla on all brains. Other sections of brains were stained when appropriate, such as those that showed high numbers of microglial nodules or infarcts. All brain sections showed no staining; however, the nasal epithelium was positive (Supplementary Figure 6; data not shown). N protein antibody staining of lung tissues of some of our patients also stained positively (data not shown).

## Discussion

Gaps in our understanding of SARS-CoV-2 infection remain. There is a paucity of detailed neuropathological data, critical to understand the neuroinvasive capacity of SARS-CoV-2 and mechanisms of neurological injury. In this retrospective study of 41 patients, we provide detailed investigations of the clinicopathological and molecular characteristics of COVID-19 in postmortem brain samples. Strengths of our study include the multi-ethnic group of patients, detailed clinic-pathological studies, the wide spectrum of hospital courses from less than a day to many weeks, and a multi-pronged effort to detect viral RNA and protein in the brains in conjunction with histopathology. Many of the pathological changes can be attributed to the effects of hypoxia, coagulopathy, and multiorgan damage in severe infection, accompanied by virus-mediated inflammatory processes, such as systemic cytokine release, while other changes reflect the age range and comorbidities of our patients.

### RNAscope and immunocytochemistry did not detect viral RNA and protein in the brains

qRT-PCR analyses of nasal epithelium and brain tissues provide evidence for the presence of viral RNA, albeit at very low levels in the brain. The high positive levels in the nasal epithelium were unexpected in some patients, given that 9 patients were sampled, at the time of autopsy, more than 1 month after their initial diagnosis by nasal pharyngeal swabs. While several RT-PCR studies have suggested prolonged shedding in some patients,^33,34^ our findings indicate that either individuals who died have prolonged viral or that viral persistence in the nasal epithelium can last even longer than previously anticipated. shedding, We detected low viral loads of SARS-CoV-2 in at least one CNS section from a substantial proportion of patients. While there is some variation among sites, the relatively low levels of viral RNA suggest that there is poor CNS tropism of SARS-CoV-2, compared to SARS or MERS. The low viral load is in concordance with recent studies,^28,32 35^ which report low viral loads in some, but not all of the brain sections tested by RT-PCR. Convincing microscopic evidence of virus in the brain is lacking in our study since RNAscope and immunohistochemistry failed to identify viral RNA or protein for the N or S regions in the brain or choroid plexus. qRT-PCR may be more sensitive, since our RNAscope detected viral RNA in lungs that had Ct values of 18-19 for the N gene (data not shown). However, a possible source of SARS-CoV-2 in brain tissues is hematogenous viral RNA within CNS blood vessels or hemorrhages, as we demonstrated in a recent case report of one of our patients (#10).^36^ It is also possible the viral RNA in some leptomeningeal vessels, as seen by RNAscope (Figure 6K), contributes to the low levels of virus seen by RT-PCR in some tissue samples. Furthermore, viral contamination during the different stages of the autopsy cannot be excluded. However, the nasal epithelium and the CNS sections were obtained during different autopsy procedures, making it unlikely that contamination from the nasal epithelium contributed to positive qRT-PCR findings. Moreover, appropriate negative controls were included in qRT-PCR assays, and repeat qRT-PCR of select sections confirmed their positive and negative status.

We interpret these very low levels of SARS-CoV-2 seen in some samples with caution. While we cannot completely exclude the possibility that viral protein or RNA is present in these brains, our observations indicate that if there, the levels must be extremely low, supporting our conclusion that the neuropathological findings are unlikely to be caused by viral infection of brain tissue. Furthermore, we found no correlations between qRT-PCR results in medullary sections and the presence of microglial activation and microglial nodules, since medullas that tested negative by qRT-PCR still contained activated microglia. Several studies have also failed to detect viral proteins by immunostaining.^28,37,38^ Rare positive cells of undefined nature have been reported^39^. Further studies examining CSF or brain from newly-infected individuals or the use of dsRNA probes to identify remnants of replicating virus may help determine any neurotropism for SARS-CoV-2.

### Microglial activation is widespread, but most common in the brainstem

A large majority of brains had prominent microglial activation in multiple areas. As expected, activation often corresponded to areas containing hypoxic damage or infarcts. However, we observed many areas of focal microglial activation characterized by microglial nodules, many of which contained neurons. These were most prevalent in the lower brainstem, although more rarely in other areas. In the medulla, both the inferior olivary nucleus and a number of tegmental nuclei were involved. Microglial nodules are characteristically present in viral and autoimmune encephalitis, in which neurons contain viral antigens or autoimmune antigens,^40^ and in animal models of recovery from neurotropic viruses, such as Zika virus, neuronophagia is observed in brain regions with demonstrated viral targeting.^41^ Despite the presence of microglial nodules and neuronophagia, we were unable to detect viral RNA or protein in these areas. Thus, it seems likely that interactions between microglia and neurons were not a direct response to viral infection of neurons, but rather were secondary to hypoxic/ischemic injury in the setting of a systemic inflammatory process.

All patients suffered some degree of hypoxia, a condition known to produce signals in neurons that attract and activate microglia,^42^ leading to phagocytosis. Microglial nodules are not commonly associated with diffuse or focal hypoxic damage, although our findings suggest that hypoxia may contribute to this type of microglial activation, particularly in the setting of a systemic inflammatory process. Furthermore, microglial activation accompanies sepsis.^43^ Although a minority of our patients were known to be bacteremic, others may have met criteria for septic shock and were receiving antibiotics. A careful review of the brains of patients with ARDS, prolonged coma, associated bacteremia, and hypoxia in pre-COVID years would be important future efforts.

An immunological cause of activated microglia and neuronophagia is a further consideration, perhaps made worse by hypoxia. The smaller microglial nodules had little or no T cell component, while larger ones included some T cells. Such pathology could represent a spectrum of inflammatory development, as postulated in Rasmussen’s encephalitis^44^, a model in which microglia first attacked neurons and secondarily attracted T cells, or T cell-mediated microglial activation and phagocytosis, as shown in flavivirus infections ^41^. Otherwise, we found little T cell presence in the parenchyma, perivascular areas, meninges, or choroid plexus.

Neuronal damage and loss in the lower brainstem could cause a number of clinical features, including altered cardiac and respiratory regulation, cranial nerve motor findings, somnolence, insomnia, and other signs and symptoms. It was unlikely that these were discovered in such ill patients, many of whom were heavily sedated on ventilatory support. The reason for a predominant brainstem localization of microglial nodules is not clear, but has been described in other reports (Supplementary Table 1). The involvement of multiple brainstem areas and nuclei are characteristic of several infectious and autoimmune disorders.^45^

### Infarcts are common

A number of our patients suffered acute or subacute infarcts, which were likely to have occurred during the course of the disease, and many of these were diagnosed postmortem. The locations of infarcts did not conform to a stereotypic pattern, since many different areas were involved in different individuals. The small infarcts associated with hemorrhages were most consistent with thrombotic or thromboembolic events and reperfusion. However, we only observed thrombi in a small number of brains (3/41; 7%). Furthermore, we did not find vasculitis, defined either as fibrinoid destruction of vessel walls or inflammatory cells within vessel walls. Indeed, immunostaining for structural components of the vascular basement membrane revealed intact blood vessels.

### Many patients have pathological co-morbidities

Given the patients’ mean age of 74, we were not surprised to see atherosclerosis and arteriolar sclerosis. These could have limited blood flow and oxygenation diffusely or focally in patients with respiratory or cardiac failure. Several brains contained cerebral amyloid angiopathy, but none was associated with hemorrhage. Some of the older patients had amyloid plaques and tau tangles and three brains contained Lewy bodies. These neurodegenerative pathologies must have preceded the SARS-CoV-2 infection, but whether or not they contributed to the acute and subacute pathologies is not clear.

### Tissue sampling in space and time

We submitted 20-30 sections per brain, including many regions. We only sampled the upper cervical spinal cord, so any lower cord pathology remains uncharacterized. Thus, even sampling many areas, we could have failed to identify relevant, local pathologies in unsampled areas.

Our patients suffered a broad time range of illness, from expiration on arrival at the hospital up to several months in the hospital (Figure 1). However, we found similar neuropathology marked by prominent microglial activation with microglial nodules and neuronophagia in patients with both short and lengthy courses. Furthermore, the fact that we did not see detectable levels of virus by RNAscope or immunohistochemistry in any of these cases, and detected only very low levels of virus by qRT-PCR, argues against the possibility that we have systematically missed some timepoint or location at which there is abundant virus after which the virus might be cleared from the brain.

### Our findings may have implications for COVID-19 survivors

It is important to consider the potential impact of the neuropathological changes we, and others, have found in autopsies if such changes are present in the brains of patients who survive COVID-19. In light of the brainstem and hippocampal distribution of microglial activation, the latter of which has been linked to virus-induced cognitive deficits,^46^ it is notable that some COVID-19 survivors develop neuropsychiatric symptoms, including memory disturbances, somnolence, fatigue and insomnia, and that similar symptoms are reported in both the acute and recovery phases. Critical to future work is understanding the short- and long-term consequences in survivors. This study included only patients who were severely ill and died. These changes may not be seen in patients with mild illness, and understanding the roles of contributing factors, such as inflammation, multiorgan damage, and hypoxia, will require further studies.

### Limitations

This study has several limitations. Our patients, a multi-ethnic group, drawn from a single center in New York City, may not represent the general COVID-19 population. Autopsy studies over-represent severe cases, and our findings may not be generalizable to less severe cases. There are currently limited data on the sensitivity and specificity of SARS-CoV-2 qRT-PCR in various tissues including brain, and positive qRT-PCR in the brain may be due to virions in the blood rather than brain tissue. Additionally, clinical data for this study were obtained through retrospective chart review and thus fully reliant on EMR. Most patients had significant pre-existing comorbidities, which confound our ability to attribute our findings to COVID-19 infection. Future studies with appropriate controls are needed to delineate which neuropathological findings result from COVID-19 infection and treatment and are not caused by other processes.

## Conclusions

1. In our single center study of 41 consecutive autopsies of COVID-19 patients we found significant neuropathology in all brains, most commonly diffuse hypoxic/ischemic damage, acute and subacute infarcts, both large and small, the latter often with a hemorrhagic component, and diffuse and focal microglial activation, including neuronophagia, predominantly localized to the brainstem. There was sparse T cell infiltration and no evidence for acute vascular wall damage. qRT-PCR on multiple frozen brain tissues of many brains showed low or absent levels of viral RNA. RNAscope and immunohistochemistry for S and N proteins were negative. Although we cannot conclusively rule out the presence of viral RNA and protein in these brains, we conclude that it is unlikely that viral infection of brain tissue directly accounts for the pathological changes.
2. Our predominantly elder, Hispanic population had multiple comorbidities including those only identified in the postmortem period. Patients died in a range of time periods, with prolonged hospital courses associated with a significant number of hospital related complications. Notably, neuropathological findings did not appear to correlate with time of hospitalization, further suggesting that pathology was not closely correlated with hospital interventions like medications or mechanical ventilation.

## Data Availability

All data is presented in the manuscript and supplementary figures and tables are included in our submission. There are no external datasets.

## Acknowledgements

The authors are most grateful to the patients’ families, who gave us consent to perform autopsies; to the many members of the autopsy staff, who performed their duties under the most difficult circumstances; to our many colleagues in the Departments of Neurology, Pathology & Cell Biology, Radiology, and Medicine, who gave their unqualified support to these studies; and to Kevin Doyle, MA, Dr. Anjali Saqi for helpful discussions and guidance with lung samples, Dr. Hanina Hibshoosh and members of the Columbia University Tissue BioBank, Dr. Jonathan Overdevest for help in removing nasal epithelium, Dr. Jean-Paul Vonsattel for his invaluable input on neuropathological diagnoses, and Ms. Trine Giaever for illustrations.

