## Supplementary Figures and Tables for "COVID-19 Neuropathology at Columbia University Irving Medical Center/New York Presbyterian Hospital"

### Supplementary Material For: COVID-19 Neuropathology at Columbia University Irving Medical Center/New York Presbyterian Hospital

#### Supplementary Figures

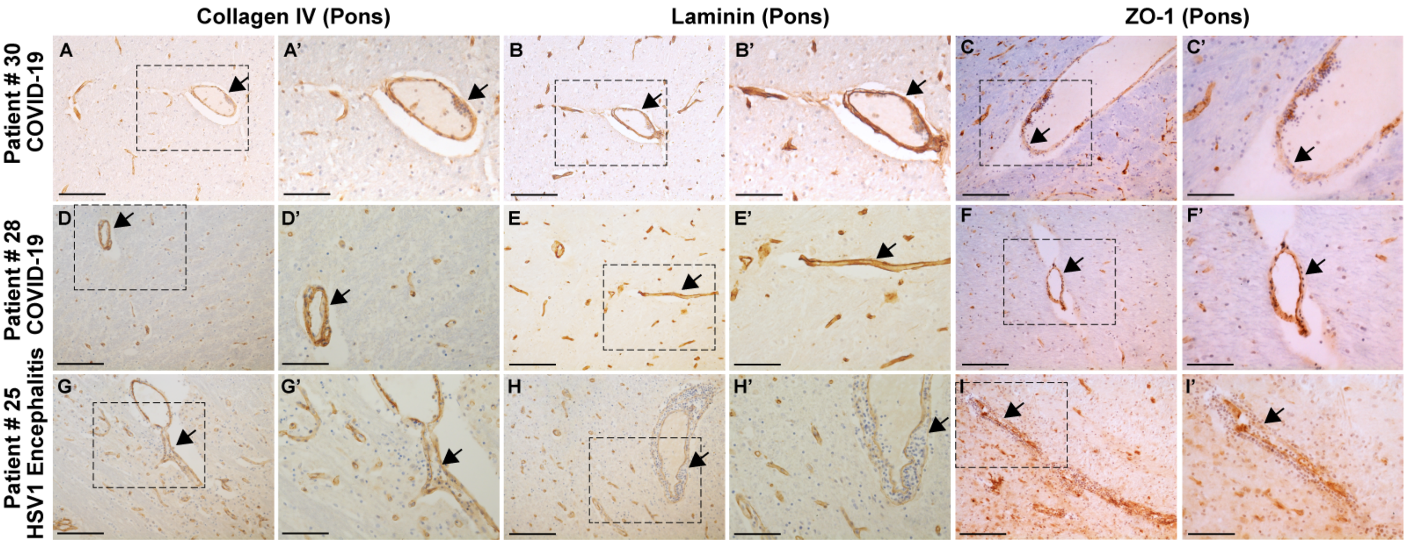


**Supplementary Figure 1. Blood vessel pathology in COVID-19 brains**. Blood vessel pathology visualized with antibodies for Collagen IV and Laminin (components of the vascular basal lamina) or on Occludens 1 (ZO-1), a tight junction protein. A-F) Representative sections from the pons of 2 COVID-19 patients show intact vascular basal lamina and tight junctions between endothelial cells. (G-I) Pontine vessels from the one patient with HSV-1 encephalitis show disrupted, separated, and poorly stained basal lamina with a large number of cellular infiltrates (G,H) or loss of tight junction staining near the cellular infiltrates (I). (E,F). A', B', C', D', E', F', G', H', I' are magnified images of boxed areas; arrows represent the same point in the corresponding images.. Scale bars: A-I 100µm, A'-I' 200µm.

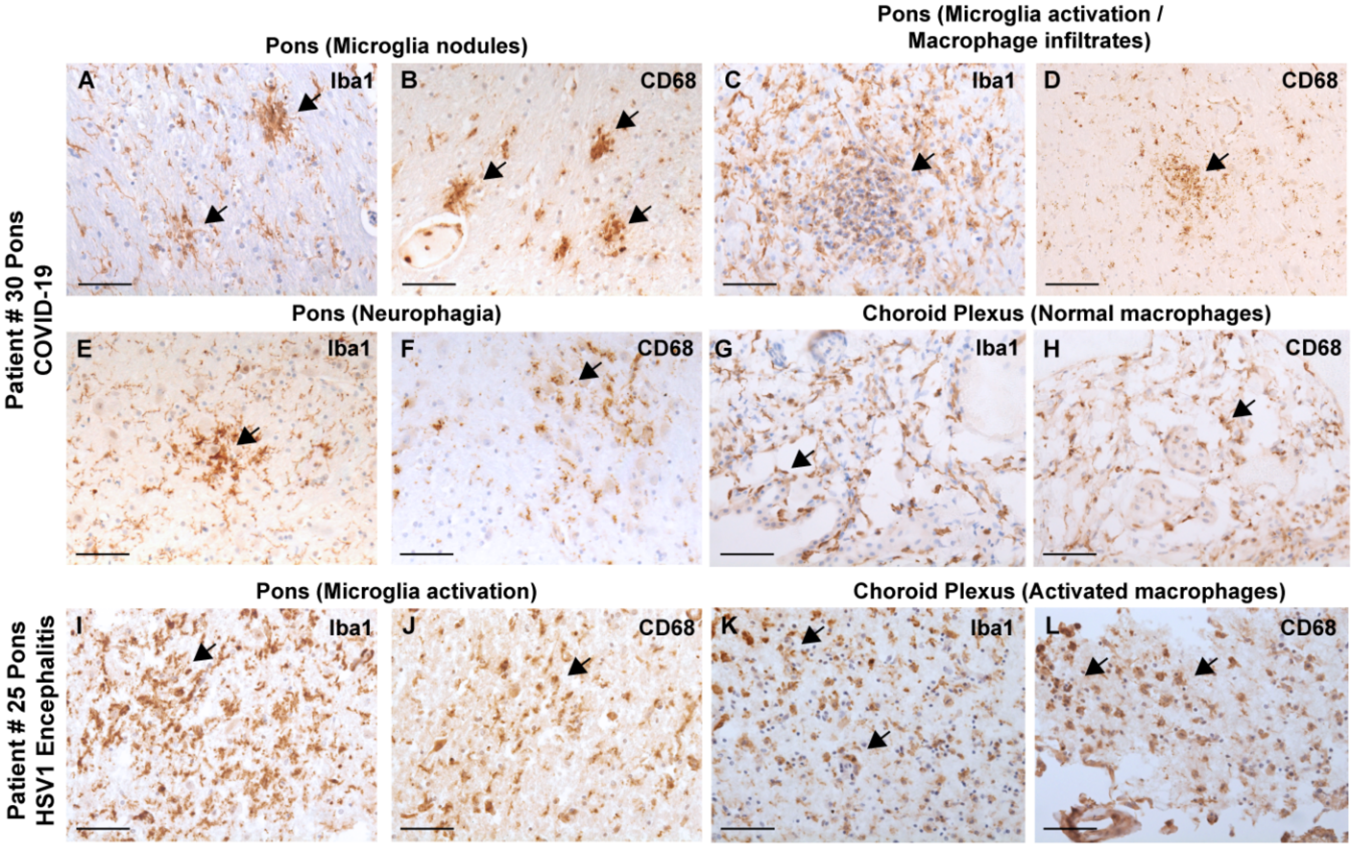


**Supplementary Figure 2. Supplementary Figure 2. Microglial pathology in the brainstem (pons) of COVID-19 cases.** Microglial pathology visualized with antibodies for Iba1 and CD68. (A-F) Representative sections from the pons of a COVID-19 patient shows examples of microglial nodules, macrophage infiltrates with some T cell involvement and neurophagia, the most prominent pathological features of the neuropathology in COVID-19 cases. G-H) Choroid plexus from the lateral ventricle stained with Iba1 and CD68 has very few macrophages. I-L) Pontine sections from the one patient with HSV-1 encephalitis shows microglia activation and macrophage infiltration into the CNS parenchyma and in the choroid plexus. Scale bars: A-L 200μm.

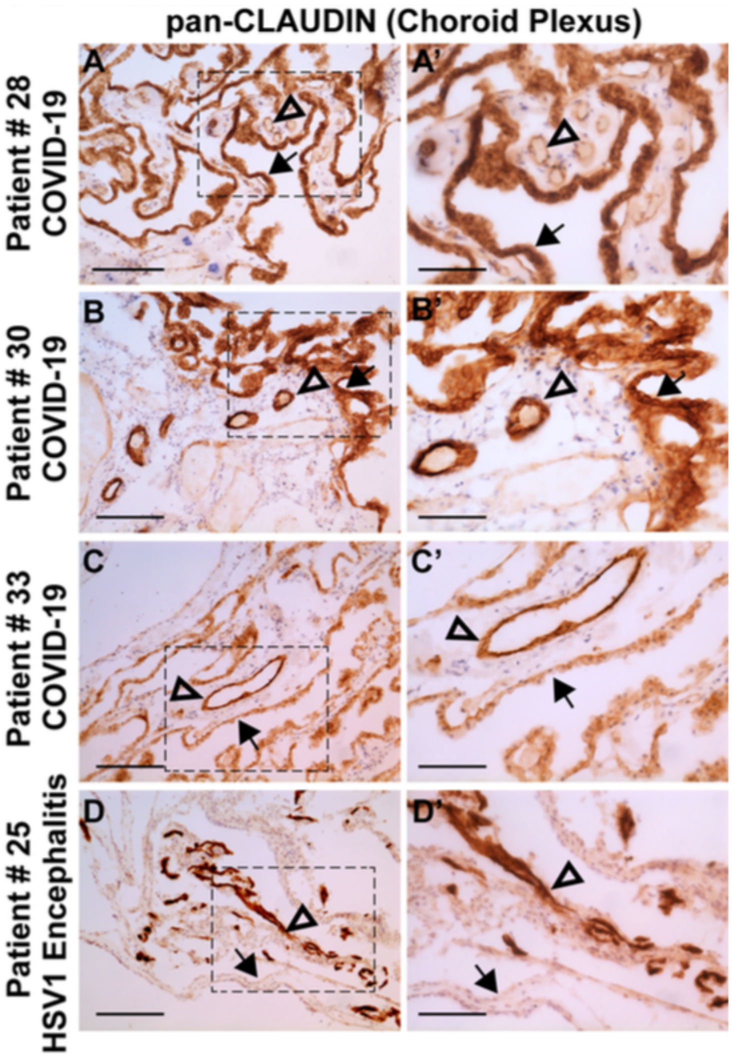


**Supplemental Figure 3. Choroid plexus epithelial barrier is intact in COVID-19 brains.** A-D') Immunohistochemistry of choroid plexus (ChP) from lateral ventricles using an antibody that recognizes both CLAUDIN-1 and CLAUDIN-5. Representative stains show expression in both the ChP epithelial cells and the ChP endothelial cells (A-C). ChP epithelium of one patient with HSV1 encephalitis shows complete loss of expression; ChP epithelia from COVID-19 patients show, to a varying extent, reduced expression. Arrows mark the ChP epithelium; open arrowheads mark the ChP endothelium. A'-D' are magnified images of boxed areas, arrows represent the same point in the corresponding images. Scale bars for A-D: 200µm. Scale bars for A'- D': 100µm.

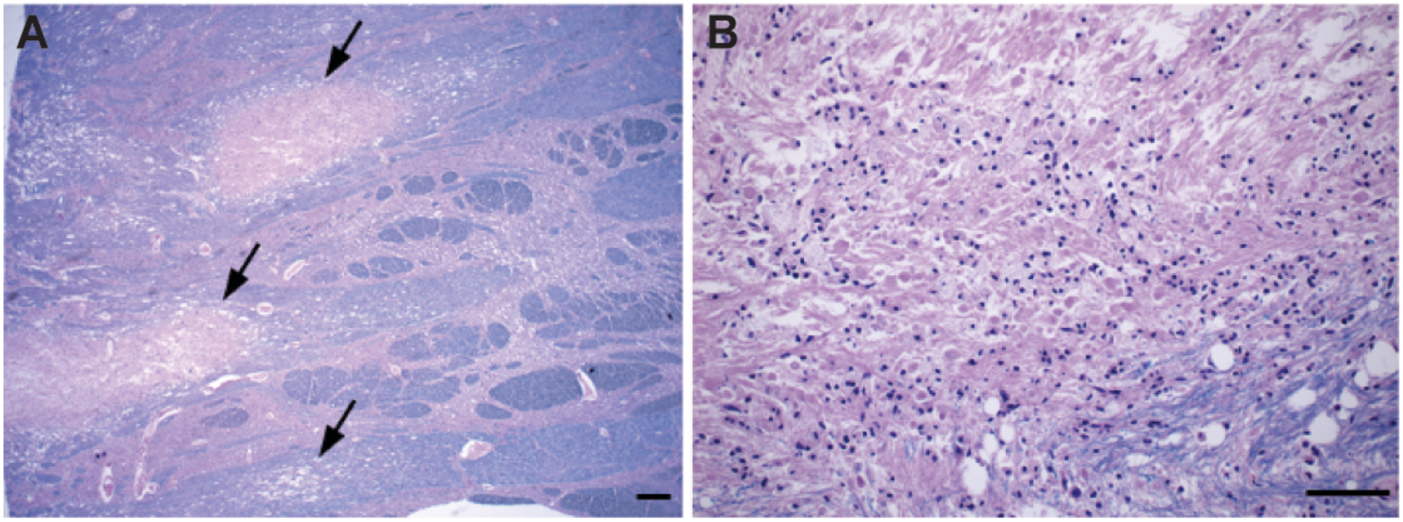


**Supplementary Figure 4. Multifocal necrotizing leukoencephalopathy in one brain.** A) Several foci of acute necrosis predominantly involving transverse fibers in the base of the pons (arrows). B) Higher magnification of a focus showing necrosis, loss of myelin, and axonal swellings. Sections are stained with Luxol fast blue stain for myelin, counterstained with H&E. Scale bars: A 1mm, B 250µm.

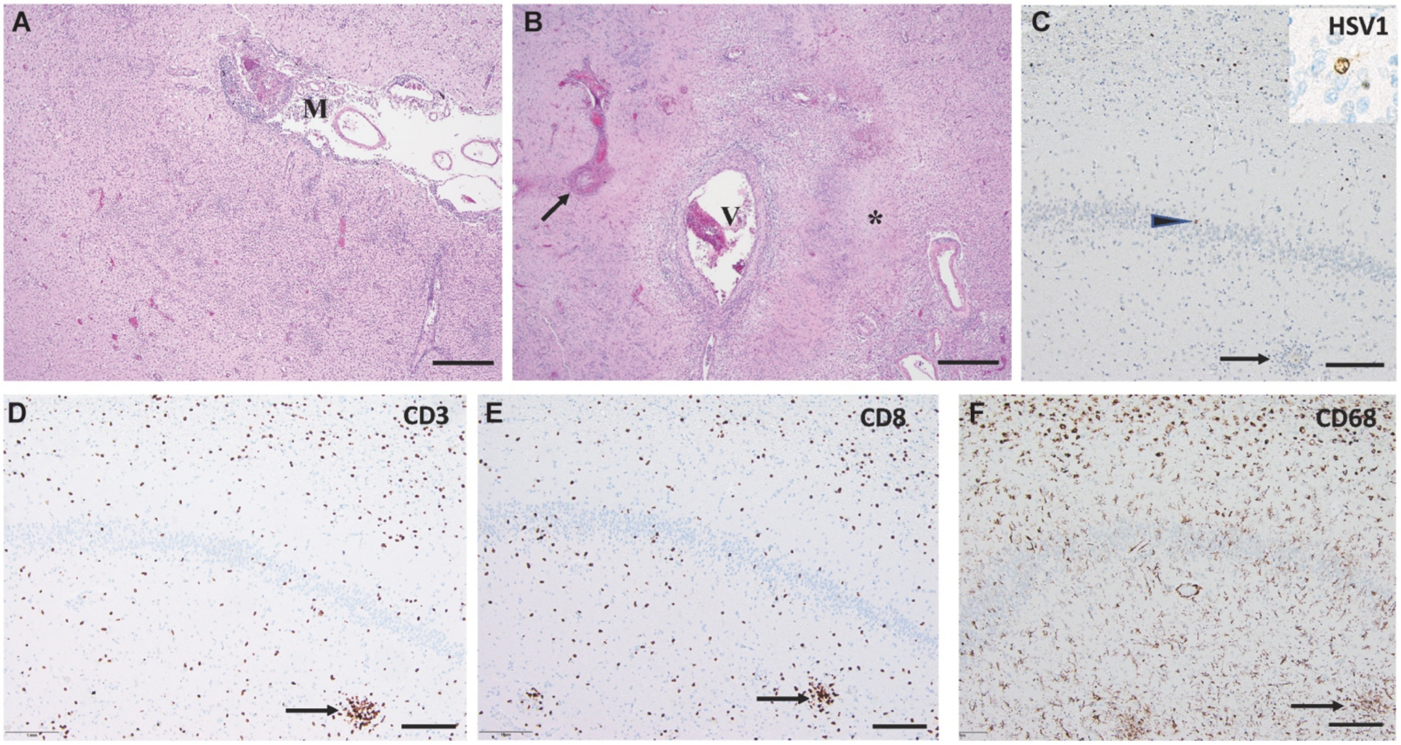


**Supplementary Figure 5. HSV-1 encephalitis in a COVID-19 patient**. A) Frontal cortex, lymphocytic infiltrates in meninges (M) and brain tissue. B) Putamen with necrosis of blood vessel wall (arrow), perivascular lymphocytic infiltrate (V), and necrosis (*). C) Hippocampus, with cell nuclei stained with HSV-1 antibody, including dentate granule neuron (arrowhead and inset) and microglial nodule (arrow). D, E, F) Hippocampus stained with antibodies for CD3, CD8, and CD68 respectively. Microglial nodules are shown by arrows. Scale bars: 1mm.

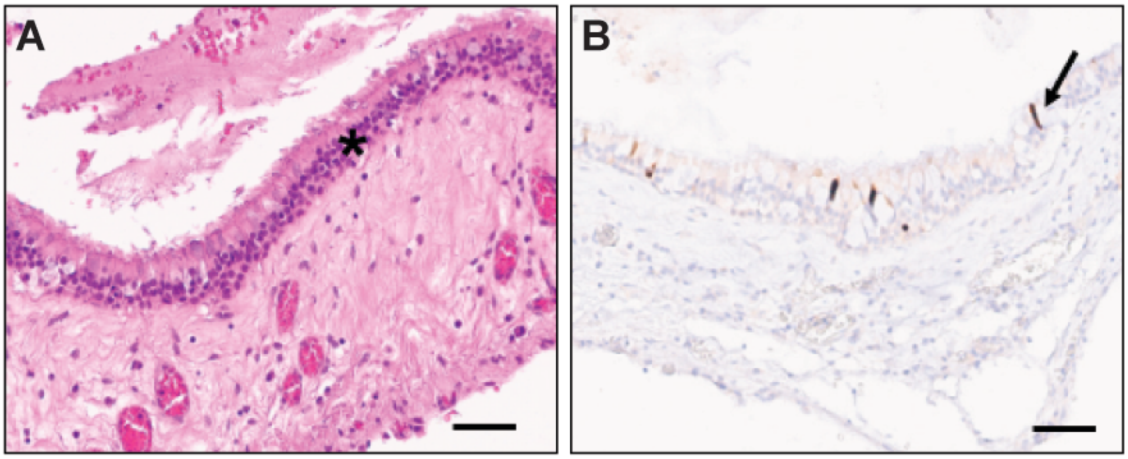


**Supplementary Figure 6. Nasal epithelium immunostained for SARS-CoV-2 N protein**. A) H&E stain or B) immunohistochemistry with the N protein antibody in the nasal epithelium (*) of a COVID- 19 patient. Several epithelial cells are positive with the N protein antibody (one marked by arrow). Scale bars: A, B 100µm.

**Supplementary Table 1.** Existing primary COVID-19 neuropathology literature.

| Authors | Study Population Location | Publication Date | Number of Patients with Brain Autopsy (%) | Histopathology | Viral and Other Tests | Gross Neuropathology | Other Findings |
|---|---|---|---|---|---|---|---|
| Meinhardt et al. [1] | Berlin, Germany | November 20 | 33 | H&E; | qRT-PCR | Not specified | Thromboembolic events 6/33 (18%) and acute infarcts. |
| Matschke *et al.* [2] | Hamburg, Germany | October 5 | 43 | H&E; GFAP, HLA-DR, TMEM119, IBA1, CD68, CD8 IHC | qRT-PCR | Six (14.0%) patients had recent ischemic infarctions. | Astrogliosis (to varying degrees) in all brains, microglia diffusely activated, with few microglial nodules in the brainstem and cerebellum. Parenchymal and perivascular $CD8^+$ cell infiltration. Cytotoxic T cells were in the frontal cortex, brainstem, and meninges. SARS-CoV-2 RNA detected in 13/27 (48.1%). |
| Hanley *et al*[3]. | London, UK | October 1 | 10 | H&E; CD3, CD20, CD4, CD8, CD68/PGM1 IHC | qRT-PCR | Varying ischemic changes in cortical neurons and white matter. One patient with large cerebral infarction with hemorrhagic transformation. | Microglial activation in 5/5 patients. Mild infiltration of T-cells observed around blood vessels and capillaries in five patients (no B cells). No necrosis or extensive inflammatory cell infiltration in parenchyma and meninges. |
| Al-Serraj *et al.* [4] | London, UK | | 8 | Histology, Immunohistochemisty | RT-PCR, *in situ* hybridization | Hemorrhagic infarction (1, 17%) | Microglial activation, few T cells, no viral RNA or protein (4, 50%), brainstem encephalitis (1, 17%), |
| Bihlmaier *et al.* [5] | Erlangen, Germany | September 15 | 3 | Immunohistochemistry (unspecified stains) | None | Two patients with white matter edema, small multifocal hemorrhages, and a bleeding patten with consecutive herniation. | No inflammatory processes in any brain. No SARS-CoV-2 RNA found in brain tissue. |

| | | | | | | | |
|---|---|---|---|---|---|---|---|
| Jensen *et al.*[6] | Cambridge, UK | September 8 | 2 | CD3, CD68 IHC | RT-PCR; RNAscope® in situ hybridization using V-nCoV2019-S probe and RT-PCR SARS-CoV-2 RNA | Cerebral cortex thinning, darkening, and calcification (Case 1). Subacute cerebellar cortex infarct (Case 2). | Brainstem encephalitis with calcifying cerebral cortical infarction and megakaryocytes. Negative ISH and RT-PCR in tissue samples of interest; specifically, no viral RNA in post-mortem brain tissue. |
| Al-Dalahmah *et al.* [7] | New York, USA | August 26 | 1 | H&E; CD3, CD8, CD68 IHC | qRT-PCR | Cerebral edema with cerebellar hemorrhage and acute infarcts in the dorsal pons and medulla. | SARS-CoV-2 RNA in the cerebellar clot, olfactory bulbs, and cerebellum. Brain sections showed severe global hypoxic changes, and several sections showed hyper-eosinophilic shrunken neurons. Microglial nodules and neuronophagia bilaterally in inferior olives and multifocally in the cerebellar dentate nuclei. Expansion of perivascular spaces in the ventral thalamus and sparce perivascular macrophages surrounding arterioles, which showed mild medial thickening. |
| Wichmann *et al.*[8] | Hamburg, Germany | August 18 | 12 | H&E | qRT-PCR | Not Specified | SARS-CoV-2 RNA was detected in the brains of four patients (33.3%). |
| Deigendesch *et al*[9]. | Basel, Switzerland | August 12 | 7 | HLA-DR, GFAP IHC | None | Not Specified | No evidence for COVID-19-related meningitis or encephalitis with increased lymphocytic infiltration of the brain or leptomeninges. Although the observed microglia activation in COVID-19 patients is a histopathological correlate of a critical illness-related encephalopathy, it was not found to be a disease-specific finding. |

| | | | | | | | |
|---|---|---|---|---|---|---|---|
| Remmelink *et al.* [10] | Brussels, Belgium | August 12 | 17 | IHC not performed on brain tissue | RT-PCR | Eight patients (47.1%) with hemorrhage, three (17.6%) with focal necrosis, five (29.4%) with edema and/or congestion, and ten (58.8%) with spongiosis. No patients demonstrated encephalitis nor vasculitis. | No patients showed isolated neuronal necrosis nor perivascular lymphocytes. Nine of eleven (81.8%) brain tissue samples had a positive RT-PCR. |
| Kantonen *et al.* [11] | Helsinki, Finland | August 6 | 4 | H&E,<br>Luxol Fast Blue,Iron | RT-PCR | One patient (25.0%) demonstrated mild brain swelling and acute microhemorrhages with enlarged perivascular spaces most pronounced in the white and deep gray matter. No meningitis nor encephalitis observed. | Three patients (75.0%) demonstrated mild-moderate hypoxic-associated changes. Few inflammatory cells found. One patient (25.0%) had few small perivascular white matter lesions and macrophages engulfing myelin. Immunostaining and RT-PCR were all negative. |
| Bradley *et al.* [12] | Washington, USA | August 1 | 5 | H&E | None | One of five (20.0%) patients demonstrated punctate subarachnoid hemorrhages and rare microhemorrhages in the brainstem. | N/A |
| Jaunmuktane *et al.* [13] | London, UK | July 8 | 2 | CD34, CD68, SMI31, and SMI94 IHC | None | One (50.0%) case with multifocal infarcts in the | One case showed dense inner rim of degenerating neutrophils and an outer rim of macrophages. The |

| | | | | | | | |
|---|---|---|---|---|---|---|---|
| | | | | | | middle and posterior cerebral arteries. One case (50.0%) with bilateral pallidal infarcts. | second case showed cortical and white matter microlesions (including hemorrhages and small infarcts), chronic infarcts, and microinfarcts in the cerebral cortex and thalamus. |
| Schaller *et al.*[14] | Augsburg, Germany | June 23 | 10 | H&E | RT-PCR | No detectable pathology. | No evidence of COVID-related encephalitis nor vasculitis. No RNA in the brain or CSF. |
| Solomon *et al.*[15] | Massachusetts, USA | June 12 | 18 | H&E; CD45, tau, amyloid-beta, and alpha-synuclein IHC | RT-PCR | Upon gross inspection, atherosclerosis was observed in 14 (77.8%) brain specimens. No acute stroke, herniation, nor olfactory bulb damage were observed. | SARS-CoV-2 was found at low levels in six brain sections of five patients. Acute hypoxic injury in the cerebrum and cerebellum. Neuronal loss in the cerebral cortex, hippocampus, and cerebellar Purkinje cell layer (no thrombi or vasculitis). In two (11.1%) brain specimens, rare foci of perivascular lymphocytes. In one (5.6%) brain specimen, focal leptomeningeal inflammation was observed. In the olfactory bulbs and tracts, no microscopic abnormalities were detected. On immunohistochemical analysis, there was no cytoplasmic viral staining. No encephalitis nor other SARS-CoV-2-specific brain changes were observed. |
| Von Weyhern *et al*[16]. | Munich, Germany | June 4 | 6 | H&E, Luxol Fast Blue; CD3 IHC | None | Neuronal cell loss and axonal degeneration were observed in the dorsal motor nuclei of the vagus nerve, trigeminal nerves, nucleus tractus solitarii, dorsal raphe nuclei, and fasciculus | Localized perivascular and interstitial encephalitis was observed. Hypoxic alterations of the brain were observed in all patients. Images showed very mild inflammatory infiltrates. Hypoxic pathology demonstrated shrunken or dying neurons and edema in many areas. |

| | | | | | | | |
|---|---|---|---|---|---|---|---|
| | | | | | | longitudinalis medialis. No territorial infarctions nor endotheliitis were observed. | |
| Efe *et al*[17] . | Samsun, Turkey | May 29 | 1 | Unspecified histopathologic examination | None | Left temporal lobe lesion. | Biopsy demonstrated perivascular lymphocytic accumulations and "hypoxic" changes. Patient was diagnosed with "encephalitis." |
| Reichard *et al.*[18] | Minnesota, USA | May 24 | 1 | H&E; CD68, APP, LFB/PAS, PLP, GFAP, CD3, and CD20 IHC | None | Lesions of likely vascular and demyelinating etiology. Hemorrhagic white matter lesions with surrounding macrophages and axonal injury. Perivascular acute disseminated encephalomyelitis (ADEM)-like lesions. | Focal necrosis with central loss of white matter and marked axonal injury. Rare neocortical organizing microscopic infarcts. |
| Duarte-Neto *et al.*[19] | São Paulo, Brazil | May 22 | 9 | H&E; TTF-1, p63, Ki67, CD4, CD8, CD20, CD57, and CD68 IHC | Ultrasound-Guided Minimally Invasive Autopsy; transsphenoidal needle puncture | Not Specified | Reactive gliosis in 8 patients (88.9%), neuronal satellitosis in 5 patients (55.6%), small vessels disease in three patients (33.3%), and perivascular hemorrhages in one patient (11.1%). |
| Paniz-Mondolfi *et al.*[20] | New York, USA | April 21 | 1 | None | Transmission Electron Microscope | Not Specified | Particles (80-110nm) spherical and pleomorphic with stalk-like projections suggestive of SARS-CoV-2 in cytoplasmic vacuoles of cells in the frontal lobe and in small vesicles of endothelium cells. Particles |

H&E = Hematoxylin and Eosin; IHC = immunohistochemistry,

| Pt # | Sex | Age Range | Race/ Ethnicity | Medical Comorbidities | Neurological Comorbidities | Presenting Neuro Symptoms | Other Presenting Symptoms | Summary of Hospital Course | Neurology consult obtained (Y/N); reason for consult; time of neuro consult/neurology admission to death | Neuroimaging Findings | Length of Mechanical Ventilation (days) | Length of hospital stay (LOHS) (days) | Length of ICU stay (days) |
|---|---|---|---|---|---|---|---|---|---|---|---|---|---|
| 1 | M | 65-70 | Hispanic | HTN, CAD | None | Impaired consciousness, weakness | Cough, SOB, myalgias, syncope collapse/agonal breathing | Found down by EMS, in ventricular fibrillation, resuscitation was attempted for 50 minutes, unsuccessful | N | | 1 | 0 | NA |
| 2 | M | 85-90 | Hispanic | Ischemic cardiomyopathy, complete heart block with PPM, interstitial lung disease with pulmonary hypertension, urothelial cancer, former smoker, DM2 | None | Weakness/fatigue, confusion, gait instability and falls | Cough, SOB, myalgias, rhinorrhea, fatigue, anorexia | He had increased work of breathing and was placed on NRB, AKI, shortly after admission cardiac arrest. 6 minutes of cardiopulmonary resuscitation and 1 round of epinephrine before death was declared. | N | Past imaging<br>MRI brain small L inferior parietal hemorrhagic infarct<br>L>R inferior frontal encephalomalacia | 0 | 1 | NA |
| 3 | M | 80-85 | Hispanic | Afib on AC, HTN. HLD, CKD, CVA, osteoporosis, depression | MCI, dementia | Confusion | None | AMS, AF w RVR, transitioned to hospice care. Text Text | N | Head CT<br>Mild MVID<br>Bilateral BG lacunar infarcts tiny<br>Head CT 18 days later, no change<br>Prior CT small right parietal infarct | 0 | 3 | NA |
| 4 | M | 70-75 | Hispanic | HTN, BPH, depression prediabetes | None | Impaired consciousness, weakness | Melena, syncope, anorexia, nausea, vomiting, fever, cough, myalgias, fatigue | Found on floor in a pool of bright red blood, hypoxic on arrival requiring NRB, hypotensive, tachycardic, worsening respiratory function-intubated. AKI, metabolic acidosis and shock, acute liver failure, coagulaopathy, GI bleed. Comfort care, extubated | N | | 4 | 7 | 5 |
| 5 | M | 55-60 | White | HTN | Unknown | None | Cough, exertional intolerance | Had arrest in the field, intubated and CPR was performed by EMS en route to ER. Ongoing efforts at resuscitation were unsuccessful in ED. | N | | 1 (intubated by EMS en route) | 0 | NA |
| 6 | M | 65-70 | Hispanic | Unknown | Unknown | None | None | Cardiac arrest during ED transport, resuscitation unsuccessful | N | | 1 (intubated by EMS en route) | 0 | NA |

| | | | | | | | | | | | | | |
|---|---|---|---|---|---|---|---|---|---|---|---|---|---|
| 7 | F | 60-65 | Hispanic | Asthma, OSA, DVT on coumadin, depression | None | Weakness/fatigue, confusion in ED (AOx2) | SOB, fever, sore throat, myalgias, anorexia | AMS, DVT, right ventricular clot, PE, AKI, hepatic dysfunction, thrombolysis and heparin drip, developed asymmetric pupils and no response to painful stimuli. Cardiac arrest en route to CT scanner | N | | 0 | 9 | 2 |
| 8 | F | 80-85 | Hispanic | COPD, CKD, HLD, osteoporosis, pelvic fractures | Dementia | Impaired consciousness | Dry cough | Hypoxic and hypernatremic on presentation, oxygen requirement worsened, AKI, transitioned to inpatient hospice. | N | | 0 | 3 | NA |
| 9 | M | 70-75 | White | Obesity, CAD s/p 5 stents, DM2 | None | None | SOB, cough, fever, nausea, diarrhea, vomiting, fatigue | NRB in ED-worsening respiratory status during admission, myocarditis, made DNR/DNI, cardiac arrest | N | | 0 | 8 | NA |
| 10 * | M | 70-75 | Hispanic | HTN, DM2 | None | Headache | SOB, abdominal pain, vomiting | Severely hypoxic and intubated by EMS, PEA arrest with ROSC 17 minutes, No brainstem reflexes-L cerebellar hemorrhage. Made comfort care. | Y: initiation of hypothermia protocol (1 day) | Head CT<br>Large R cerebellar hemorrhage with intraventricular extension, surrounding edema in cerebellum, pons and medulla<br>SAH in posterior fossa, basal cisterns and r MCA cistern<br>Hydrocephalus and loss of grey white differentiation concerning for hypoxic injury<br>CTA negative<br>Prior MRI IAC mild MVID | 1 (intubated at nursing home before admission) | 1 | NA |
| 11 | F | 35-40 | Hispanic | Post-liver transplant 2017, TB | Hepatic encephalopathy | None | Diarrhea, abdominal pain, jaundice, dark urine | Admitted for allograft rejection, developed leukocytosis in hospital-COVID 19 positive. Hypoxemic respiratory failure, fungemia , AKI, coagulopathy , pancytopenia and AMS. Hospital day 27 non-reactive pupils, Multifocal ICH, global edema, herniation. Brain death. | Y: ICH found on Head CT in setting of persistent AMS (2 days) | Head CT<br>Large Right temporal hemorrhage, extensive SAH, focal small parenchymal hemorrhages bilateral frontal, parietal , temporal lobes and cerebellar hemispheres –possible embolic infarcts<br>Effacement of cisterns and loss of gray white c/w severe hypoxemic | 13 | 28 | 13 |

| | | | | | | | | | | | | | |
|---|---|---|---|---|---|---|---|---|---|---|---|---|---|
| | | | | | | | | | | injury and cerebral edema<br>Head CT WNL<br>Prior head 3 years before WNL | | | |
| 12 | F | 75-80 | Hispanic | DM2, HTN, congenital solitary kidney | None | None | Cough, fever, SOB, chills | NRB in ED, intubated, AKI, CVVH, pneumothorax, chest tube and Gill procedure, ischemic left upper extremity started on AC, bacteremia, persistent elevated oxygen requirements, transitioned to comfort care and compassionately extubated. | N | | 12 | 19 | 11 |
| 13 | F | 75-80 | Hispanic | Asthma, CAD, CVA, PAD, CKD | Stroke | None | Cough, SOB, fatigue | NC and NRB in ED, AKI, DNR/DNI, transitioned to comfort care. | N | | 0 | 15 | NA |
| 14 | M | 75-80 | Hispanic | HTN, CAD, COPD | Dementia | Confusion, impaired consciousness, combativeness | Lethargy, poor PO intake | AMS, AKI, hyponatremia on presentation, agitated delirium, intestinal ileus, witnessed aspiration event, made comfort care. | N | | 0 | 17 | NA |
| 15 | M | 60-65 | Hispanic | HTN | None | Headache | Fever, SOB, fatigue, diarrhea | In ED on NRB, respiratory status worsened, intubated , developed ARDS, ARF requiring CRRT, worsening lactatemia, anion gap metabolic acidosis and shock liver. Hospital day 13, cardiac arrest. | N | | 3 | 13 | 3 |
| 16 | M | 80-85 | Hispanic | HTN | PD | Confusion | SOB, chest tightness | NRB in ED, UTI, Day 8, oxygen requirement increased and made comfort care. | N | | 0 | 9 | NA |
| 17 | M | 75-80 | Black | HTN, HLD, DM2, CAD, HFrEF, CKD | Dementia | Confusion | Fever, cough, SOB, poor PO intake | AMS, Acute on chronic renal failure. Worsening respiratory status, made DNR/DNI. Oliguric renal function, decision not to pursue dialysis. Transferred to inpatient hospice. | N | | 0 | 11 | NA |
| 18 | M | 80-85 | Hispanic | Bedbound with joint contractures, schizophrenia, prostate cancer | Seizure disorder | Confusion, impaired consciousness | SOB, hypoxia | AKI and hypernatremia on arrival to ED, made DNR. Intubated, started on vasopressors, urosepsis, atrial fibrillation with RVR, extubated, though mental status remained poor, transitioned to hospice care. | N | | 10 | 14 | 10 |
| 19 | F | 95-100 | Hispanic | HTN, HL, CAD, CVA, SVT, Depression | Dementia, stroke | Confusion | None | In ED became more lethargic, severe hypoxemia and bradycardia. DNR/DNI, transitioned to comfort care, and died in the ED. | N | No images in PACS<br>No CXR | 0 | 0 | NA |

| | | | | | | | | | | | | | |
|---|---|---|---|---|---|---|---|---|---|---|---|---|---|
| 20 | F | 65-70 | Hispa nic | DM2, HTN, former cigar smoker | None | Weakness | SOB, fever | Intubated in ED, AKI, CVVH, bacterial pneumonia. Progressive multiorgan failure, made DNR, no escalation of therapy. | N | | 25 | 25 | 21 |
| 21 | M | 65-70 | Hispa nic | HLD, HTN, past prostate cancer | None | Back pain | SOB, fever, fatigue | AMS, DVT, DKA, intubated for hypoxic respiratory failure, new seizures and embolic infarcts. Transferred to hospice. | Y: Multifocal acute infarcts found on Head CT, new onset seizure (1 day) | Head CT and CTA Acute infarcts R basal ganglia, Right inferior frontal lobe and left superior frontal/ parietal acute infarcts possibly embolic CTA no large vessel occlusion AUTOPSY BRAIN Right inferior frontal, right basal ganglia and left frontal parietal subacute infarcts all with associated hemorrhage | 21 | 26 | 21 |
| 22 | F | 70-75 | Hispa nic | HLD, hypothyroidism | None | None | SOB, fever, chills, cough, myalgias | Intubated during hospitalization, ARDS, AKI, CRRT, bacterial pneumonia, UTI. Tracheostomy, not awakening despite sedation wean, worsening mental status. | N | Head CT Mild MVID, small l> R basal ganglia old lacunar infarcts AUTOPSY right parietal signal abnormality possible recent WM infarct, suggestion of cortical hemorrhage ( side of largest brain cuts) | 39 | 39 | 38 |
| 23 | M | 70-75 | Hispa nic | ESRD on HD, CAD s/p CABG, HFpEF, pHTN, HTN, HLD, DM2, OSA, CKD | None | Impaired consciousness | None | Cardiac arrest at home, ROSC 15 minutes, refractory shock, global cerebral edema | N | Head CT Extensive loss of grey white matter c/w severe hypoxemic injury | 0 | 0 | 0 |
| 24 | M | 65-70 | Hispa nic | HTN, HLD, DM | None | Weakness | Fever, cough, sore throat, SOB, fatigue, anorexia, rhinorrhea, diarrhea | Respiratory failure, intubated in hospital, course c/b ICH, seizures, AKI, CVVH, made comfort care | Y: SDH found on Head CT, frontal mass and SAH found on Brain MRI, new onset seizure (17 days) | Head CT Left frontal parenchymal hemorrhagic lesion likely subacute infarct with moderate surrounding edema right frontal small acute subdural hemorrhage and diffuse SAH head Ct later same day no change | 41 | 46 | 41 |

| | | | | | | | | | | | | | |
|---|---|---|---|---|---|---|---|---|---|---|---|---|---|
| | | | | | | | | | | brain MRI<br>Diffuse hypoxemic, hypoperfusion injury<br>Punctate hemorrhages throughout the bilateral basal ganglia<br>And bilateral pons with few scattered inferior frontal lobes and left parietal lobe | | | |
| 25 | F | 65-70 | Hispanic | HTN | None | Weakness | Fever, SOB | NRB in ED, intubated. ARDS. AKI, bacteremia, VAP, CMV viremia, UTI. Palliatively extubated in context of refractory hypoxemia. | N | | 42 | 42 | 41 |
| 26 | M | 70-75 | Hispanic | HTN, history of colon cancer | None | None | Fever, cough, SOB, myalgias, fatigue, abdominal pain | ARDS, intubated, bacterial pneumonia, candidemia, acidemia, suspected PE, AKI, GI bleeding. terminally extubated. | N | | 21 | 29 | 21 |
| 27 | M | 75-80 | Hispanic | HTN, former smoker, CKD s/p nephrectomy, AF on home AC | None | None | SOB, chills | Prior two week admission for COVID, on NRB during admission but not intubated. Possible bacterial pneumonia, AKI, DVT, worsening respiratory status, AMS, cardiac arrest | N | Head CT<br>Mild MVID small superior bifrontal lobe infarcts unhanged<br>Medial L.R temporal lobe hypodensities , possible subacute infarcts | 0 | 5 | NA |
| 28 | F | 75-80 | Hispanic | HTN, DM2, renal transplant, CAD, depression | None | Confusion, impaired consciousness | SOB, fatigue, chest pain, abdominal pain | Hypotensive requiring pressors, DVT, AKI. DNR/DNI. | N | | 0 | 2 | 1 |
| 29 | M | 60-65 | Hispanic | HTN, CKD, ILD s/p lung transplant, chronic strongyloides infection | PD | None | SOB | Suspected PE, worsening grant involvement, bacterial pneumonia, AKI, CVVH, ARDS, pneumothorax. Terminally extubated. | N | Prior brain MRI 4 years ago Mild MVID, old Right frontal, Left insula, Left superior temporal infarcts | 7 | 22 | 10 |
| 30 | M | 60-65 | Hispanic | None | None | None | Fever, cough, SOB, chills, sore throat, chest pain | NRB in ED intubation, ARDs, tension pneumothorax with chest tubes, diffuse subcutaneous emphysema, VAP, tracheostomy, AKI, DVT, Persistent pressor requirement. | N | | 33 | 35 | 33 |
| 31 | F | 65-70 | Declined | HTN, DM2, hypothyroidism | None | Anosmia, Ageusia | Fever, cough, SOB, myalgias | NRB, intubated, ARDS, bacterial pneumonia, AKI, right facial droop. Initial clinical improvement, then persistent | N | AUTOPSY<br>Possible punctate BG hemorrhage | 18 | 38 | 35 |

| | | | | | | | | | | | | |
|---|---|---|---|---|---|---|---|---|---|---|---|---|
| | | | | | | | | tongue swelling and airway edema, treated by allergy. R facial droop noted two days before death, cardiac arrest. | | | | |
| 32 | M | 70-75 | Hispanic | HTN, HLD, DM2, BPH | None | None | Fever, cough, SOB, sore throat, chills, myalgias | NRB in ED, intubated. ARDS, pneumothorax with chest tube placement, subcutaneous emphysema. Bacterial pneumonia, AKI, CVVH, diabetic ketoacidosis requiring insulin drip, anemia requiring multiple blood transfusions, hypernatremia and vasodilatory shock, tracheostomy with persistent oxygen requirements, transitioned to comfort care. | N | AUTOPSY<br>Possibly right occipital horn hemorrhage | 45 | 48 | 45 |
| 33 | M | 65-70 | Hispanic | CKD | Dementia, PD | Impaired consciousness, confusion | Fever, SOB | Intubated on admission, septic shock, bacterial pneumonia, AKI, CVVH, tracheostomy placed, initial improvement in respiratory status. Persistent agitated delirium. Transitioned to comfort care. | Y: Parkinson's medication initiation in setting of AMS (17 days) | | 31 | 39 | 38 |
| 34 | M | 90-95 | Hispanic | HTN, BPH | None | Impaired consciousness, confusion, weakness | Anorexia | Rhabdomyolysis, SARS-Cov2 PCR positive during admission, UTI, bacteremia, necrotic sacral decubitus ulcer, atrial fibrillation with RVR, non-ST elevation MI, worsening heart failure, PHTN. Found unresponsive, pulseless. No resuscitation per family wishes. | Y: dysarthria (46 days) | Head CT and MRI stroke protocol<br>moderate MVID<br>no acute infarct , no hemorrhage L parietal tiny WM infarct<br>AUTOPSY<br>Left cerebellar infarcts on T2<br>(T2 not obtained in vivo) | 0 | 48 | NA |
| 35 | F | 85-90 | Hispanic | HTN, HLD, PPM, AF, iron deficiency anemia | Dementia, stroke | Decreased consciousness, confusion | None | On arrival patient was unresponsive, stroke code activated. Head CTA possible hyperdense basilar artery, VAP, comfort care. | Y: concern for stroke (5 days) | | 0 | 5 | NA |
| 36 | M | 75-80 | White | Waldenstrom macroglobulinemia, Asthma, OSA, COPD | None | None | Fever, cough, SOB | ARDS, AKI, CRRT, pneumothorax. Transitioned to comfort care 10 weeks after initial presentation. | N | AUTOPSY<br>No significant abnormality | 68 | 60 | 60 |
| 37 | M | 55-60 | Unknown | stroke | None | None | SOB | Cardiac arrest during ED transport, unsuccessful attempts of intubation by EMS, supraglottic airway was placed. Pulseless on arrival to the ED, died in ED. | N | | 0 | 0 | NA |

| | | | | | | | | | | | | | |
|---|---|---|---|---|---|---|---|---|---|---|---|---|---|
| 38 | M | 70-75 | Hispanic | HTN, CAD, CKD, s/p renal transplant | None | None | Cough, SOB, diarrhea, abdominal pain | AKI, Shigella colitis, progressive hypoxemic respiratory failure. Empiric therapeutic anticoagulation. CMV viremia, received ganciclovir, transferred to ICU for worsening respiratory status and made comfort care 33 days after admission. | N | AUTOPSY<br>Bilateral occipital small intraventricular hemorrhage<br>Parietal parenchymal possible small hemorrhage<br>Superior white matter infarct /MVID | 0 | 33 | 15 |
| 39 | F | 72-75 | Hispanic | HTN | None | None | Cough, rhinorrhea | Intubated in ED, AKI, VAP, unable to wean from ventilatory support, transitioned to comfort care 68 days after admission. | N | AUTOPSY possible intraventricular temporal horn hemorrhage<br>Suggestion of Tiny parietal hemorrhage | 68 | 69 | 68 |
| 40 | F | 85-90 | Hispanic | HLD, HTN, DM, PVD, depression | Dementia, L MCA stroke | Decreased consciousness, confusion | Nausea/vomiting, anorexia, abdominal pain | Hypotensive in ED, transitioned to comfort care. | N | | 0 | 1 | 1 |
| 41 | M | 70-75 | Declined | HTN, obesity, former smoker | None | Back pain, right leg weakness, sensory symptoms | None | Developed sudden onset tearing back pain and right leg weakness with CT angiogram showing aortic dissection extending from the aortic root to the infrarenal aorta with hemopericardium. Tested positive for COVID 19, not a surgical candidate. AKI, PE, AMS. Started on anticoagulation, cardiac arrest with no ROSC achieved until he was cannulated to VA_ECMO after 26 minutes of ischemic time. Intubated, targeted temperature management with mental status remained poor prompting head CTA global anoxic brain injury. Transitioned to comfort care 22 days after initial presentation. | Y: initiation of hypothermia protocol (9 days) | Head CT<br>Moderate MVID<br>Left thalamic, Left anterior limb IC<br>Left pons small infarcts indeterminate age<br>Acute left occipital lobe infarct<br>Head CT<br>No change<br>Basilar artery appears slightly dense, possible thrombus<br>As above plus loss of grey white slight, early hypoxemic injury | 10 | 22 | 10 |

OSA=obstructive sleep apnea. DVT=deep venous thrombosis. PE=pulmonary embolism. AMS=altered mental status. HTN=hypertension. HLD=hyperlipidemia. DM2=Diabetes Mellitus type 2. SOB=Shortness of breath. EMS=Emergency Medical Services. PEA=Pulseless electrical activity. ROSC=Return of spontaneous circulation. PPM=Permanent pacemaker. NRB=nonrebreather mask. CAD=Coronary artery disease. DNR=Do not resuscitate. DNI=Do not intubate. CRRT=Continuous renal replacement therapy. CVVT=Continuous veno-venous dialysis. UTI=Urinary tract infection. CRF=Chronic renal failure. ARF=Acute renal failure. AF=Atrial fibrillation. RVR=Rapid ventricular rate. MCA=Middle cerebral artery. PD=Parkinson's Disease. PVD=Peripheral vascular disease. PHTN=Pulmonary hypertension. DKA=Diabetic ketoacidosis. ECMO=Extracorporeal membrane oxygenation. NA=Not applicable. *Patient published (Al Dalahmah *et al.*, 2020)

**Supplementary Table2.** Summary of cases: Patient Characteristics including neuroimaging findings

#### Supplementary Table 3. Laboratory Values

| Pt # | WBC on admission [x(10^3)/uL] | Peak WBC [(10^3)/uL] | Peak CRP [mg/L] | Peak ESR [mm/hr] | Peak LDH [U/L] | Peak Ferritin [ng/mL] | Peak IL-6 [pg/mL] | Peak D-dimer [ug/mL] | Peak Fibrinogen [mg/dL] | Peak Glucose [mg/dL] | Peak Creatine Kinase[U/L] | Peak Creatinine [mg/dL] | Peak Lactate [mmol/L] | Peak Procalcitonin [ng/mL] |
|---|---|---|---|---|---|---|---|---|---|---|---|---|---|---|
| Patient numbers | 3.48 - 9.42 | 3.48 - 9.42 | 0.00 - 10.0 | 0 - 20 | 135-214 | 13.0 - 150.0 | < = 5.0 | 0.00 - 0.80 | 191 – 430 | 75 - 100 | 40.0 - 308.0 | 0.50 - 0.95 | 0.5 - 2.2 | < = 0.08 |
| 1 | | | | | | | | | | | | | | |
| 2 | 5.7 | 6.8 | | | | | | | 14.1 | 335 | | 1.6 | 3.4 | 0.16 |
| 3 | 3.4 | 10.5 | | 46 | | | | | | 131 | | | 3.5 | |
| 4 | 5.1 | 14.0 | 297.1 | 63 | 1119 | 9897 | >315.0 | >20.00 | 107 | 415 | 3074 | 4.5 | 9.1 | 45.95 |
| 5 | | | | | | | | | | 115 | | | | |
| 6 | | | | | | | | | | | | 13.49 | | |
| 7 | 29.9 | 40.9 | 300 | 130 | 4305 | 3623 | 66.1 | >20.00 | >700 | 265 | 823 | 2.5 | 2.1 | 7.38 |
| 8 | 12.7 | 12.7 | 30.3 | 53 | 417 | 762.8 | | | | 114 | 287 | 2.2 | 3.6 | 0.24 |
| 9 | 24.1 | 24.1 | 211.8 | 56 | 1406 | 4460 | >315.0 | >20.00 | | 285 | | 1.2 | 5.4 | 0.46 |
| 10 | 5.8 | 13.3 | 8.7 | 27 | | 38 | | | | 462 | | 2.7 | 8.4 | |
| 11 | 6.9 | 17.5 | 155 | 66 | 1336 | 5868 | >315.0 | 19.14 | 662 | 392 | 162 | 3.48 | 3.2 | 9.61 |
| 12 | 5.1 | 22.0 | 300 | 130 | 591 | 1056 | >315.0 | >20.00 | | 528 | 199 | 6 | 3.8 | 6.63 |
| 13 | 6.8 | 25.1 | 205.59 | 117 | 512 | 391.8 | 248 | 9.44 | 205.6 | 172 | 71 | 1.4 | 3.8 | 0.25 |
| 14 | 22.4 | 25.3 | 98.4 | 28 | | 730.8 | | | | 159 | 974 | 3.53 | 5.1 | 0.37 |
| 15 | 25.3 | 49.9 | 300 | 105 | 5000 | 100000 | >315.0 | >20.00 | 415 | >571 | 783 | 3.22 | 16 | 3.19 |
| 16 | 7.3 | 23.5 | 123.9 | 35 | 642 | 1181 | 206.8 | 1.01 | 461 | 208 | 675 | 1.14 | 1.7 | 16.3 |

| | | | | | | | | | | | | | | |
|---|---|---|---|---|---|---|---|---|---|---|---|---|---|---|
| 17 | 9.7 | 15.1 | 205.9 | 102 | 1058 | 2171 | 135.4 | 11.99 | 697 | 182 | 434 | | 1.5 | 1.47 |
| 18 | 13.6 | 40.9 | 285.2 | 126 | 810 | 505.8 | >315.0 | 9.56 | | 321 | 2486 | 2.87 | 4.6 | 36.09 |
| 19 | 1.4 | 4.8 | | | 352 | 3194 | | | | | | | 3.1 | 0.11 |
| 20 | 10.8 | 50.9 | 300 | 80 | 549 | 1992 | >315.0 | >20.00 | | 371 | 566 | 6.83 | 3.7 | 6.98 |
| 21 | 5.4 | 19.7 | 277.83 | 63 | 955 | 1003 | >315.0 | >20.00 | | 286 | 796 | 1.91 | 2.9 | 1.69 |
| 22 | 17.35 | 20.9 | 300 | 130 | 887 | 1341 | >315.0 | 6.04 | 564 | 241 | 704 | 6.12 | 4.2 | 15.39 |
| 23 | 6.3 | 15.2 | 156.5 | 115 | 4538 | 39616 | 8.3 | 13.49 | | 130 | 119 | | 11.8 | 15.1 |
| 24 | 9.7 | 19.4 | 300 | 112 | 1647 | 3896 | >315.0 | >20.00 | | | | | 2.2 | 20.6 |
| 25 | 15.67 | 24.73 | 300 | 115 | 601 | 1160 | >315.0 | >20.00 | | 180 | 154 | 1.95 | 2.4 | 60.45 |
| 26 | 15.95 | 35.3 | 300 | 65 | 626 | 2371 | >315.0 | 16.05 | 349 | 163 | 5293 | 0.84 | 3.2 | 0.55 |
| 27 | 19.5 | 19.5 | 373.06 | 130 | 638 | 1639 | | >20.00 | >700 | 219 | 31 | 2.21 | 2.1 | 1.38 |
| 28 | 13.9 | 13.9 | | | 291 | 1213 | | | | 419 | 136 | 1.26 | 3.5 | 18.1 |
| 29 | 31.6 | 40.6 | 233.93 | 39 | 1730 | 2853 | >315.0 | >20.00 | 667 | 320 | 612 | 3.21 | 4.5 | 0.92 |
| 30 | 9.6 | 30.4 | 300 | 118 | 1083 | 1282 | >315.0 | >20.00 | 877 | 267 | 2522 | 3.97 | 2.5 | 31.3 |
| 31 | 11.6 | 14.7 | 202.64 | 80 | 865 | 2465 | 76.6 | >20.00 | 296 | 422 | 3950 | 1.22 | 4.3 | 1.07 |
| 32 | 11.2 | 30.8 | 300 | 100 | 397 | 2995 | >315.0 | >20.00 | 1053 | 398 | 838 | 2.69 | 6.2 | 5.19 |
| 33 | 11.03 | 21.5 | 300 | 130 | 517 | 2395 | >315.0 | >20.00 | 411 | 160 | 645 | 6.56 | 2.1 | 25.6 |
| 34 | 7.3 | 23.5 | 223.65 | 130 | 574 | 2458 | 120.6 | 11.39 | | 160 | 13074 | 2.29 | 1.4 | 0.28 |
| 35 | 8.9 | 8.9 | 0.44 | 10 | | | | 1.23 | | 136 | 61 | 1.06 | 1.2 | 0.81 |
| 36 | 11.18 | 40.8 | 12.87 | 130 | 380 | 2823 | >315.0 | >20 | 613 | 226 | 49 | 8.66 | 2.1 | 7.19 |
| 37 | | | | | | | | | | | | | | |
| 38 | 6.4 | 31.2 | 300 | 94 | 681 | 1877 | >315.0 | 3.95 | 548 | 583 | 786 | 1.95 | 4.9 | 2.15 |
| 39 | 26.6 | 33.1 | 237.2 | 130 | 773 | 1988 | >315.0 | 14.39 | 529 | 5782 | 684 | 4.52 | 3.1 | 6.1 |
| 40 | 15.6 | 15.6 | | | | | | | | 220 | | 0.95 | 10 | |

| | | | | | | | | | | | | | | |
|---|---|---|---|---|---|---|---|---|---|---|---|---|---|---|
| 41 | 13.6 | 14.8 | 213.98 | 73 | 1599 | 516.7 | 74.4 | >20.00 | 507 | 181 | 1743 | 6.43 | 4.1 | 65.6 |

Supplemental Table: Laboratory values

**Supplementary Table 4: SARS-CoV-2 is Detected in Different Areas of the Brain by qRT-PCR**

| Brain Autopsy Section (n=) | Number of samples positive for SARS-CoV-2 (%) | Median viral copy/sample (IQR) |
|---|---|---|
| Nasal Epithelium (21) | 19 (91%) | 43,840 (99,360) |
| Olfactory Bulb (25) | 10 (40%) | 680 (256) |
| Temporal Lobe (25) | 9 (36%) | 928 (416) |
| Cerebellum (23) | 10 (44%) | 264 (216) |
| Medulla (24) | 8 (33%) | 1440 (272) |
| Superior Frontal Gyrus (7) | 1 (14%) | 368 (0) |

**Supplementary Table 5: RNAscope data from fresh frozen brains of COVID-19 cases**

| Case # | Region | RT-PCR | RNAscope | Case # | Region | RT-PCR | RNAscope |
|---|---|---|---|---|---|---|---|
| 1 | OB<br>ME | High (+)<br>Low (+) | Negative<br>Negative | 16 | ME | Positive | Negative |
| 3 | ME | Low (+) | Negative | 18 | ME | Low (+) | Negative |
| 5 | ME | Low (+) | Negative | 21 | ME | Positive | Negative |
| 6 | OB<br>ME | Positive<br>Positive | Negative<br>Negative | 22 | ME | Negative | Negative |
| 7 | ME | Indeterminate | Negative | 24 | ME | Negative | Negative |
| 9 | ME | Low (+) | Negative | 36 | ME | ND | Negative |
| 11 | OB<br>CE | Positive<br>Positive | Negative<br>Negative | 37 | ME | ND | Negative |
| 12 | ME | Negative | Negative | 40 | ME | ND | Negative |

OB: Olfactory bulb

ME: Medulla

CE: Cerebellum

ND: Not Determined

Low (+): Low positive

**Graphical Abstract**

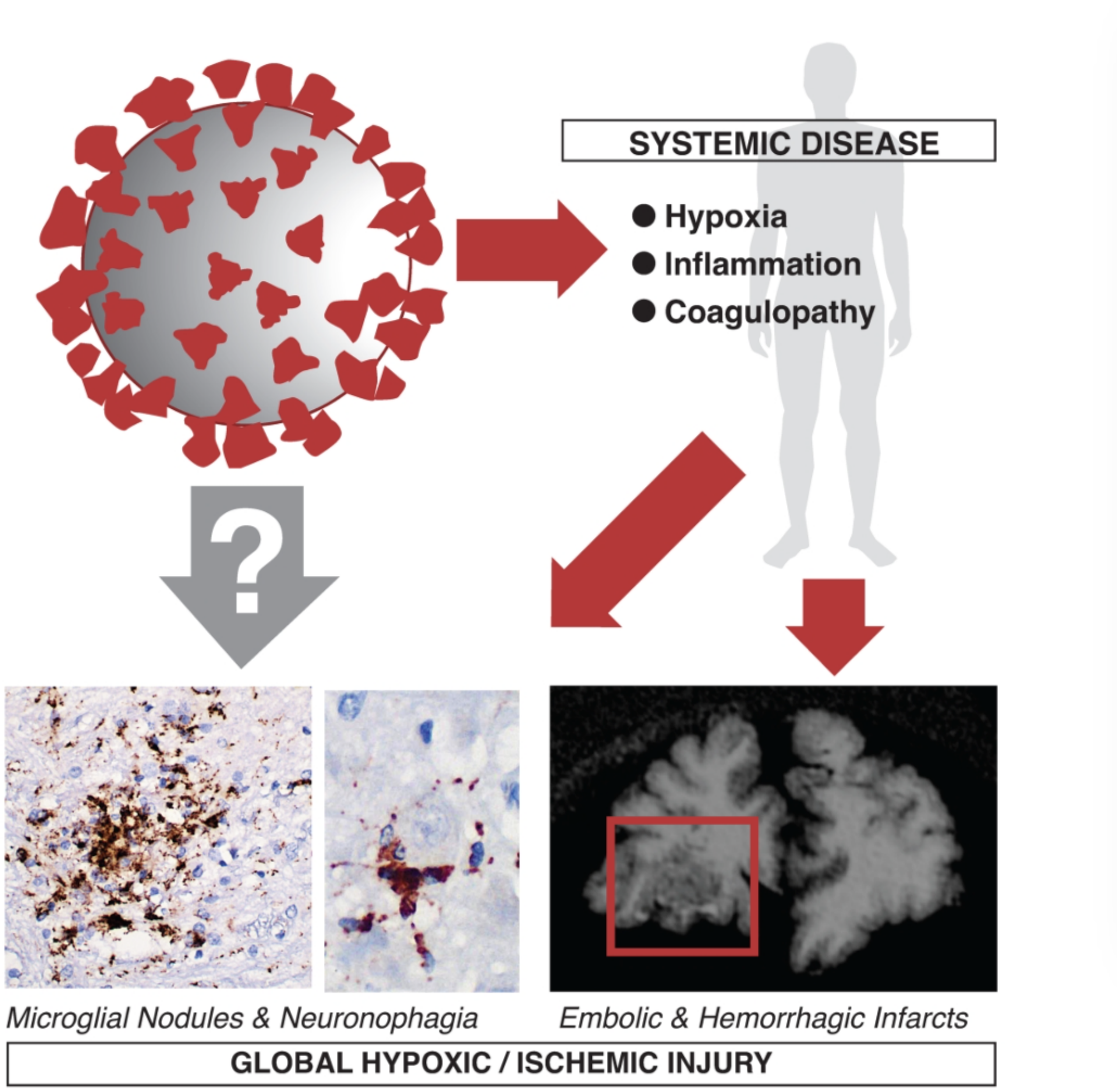
